# Measuring Voluntary Responses in Healthcare Utilization During the COVID-19 Pandemic: Evidence from Taiwan

**DOI:** 10.1101/2020.11.28.20240333

**Authors:** Yung-Yu Tsai, Tzu-Ting Yang

## Abstract

Healthcare has been one of the most affected sectors during the coronavirus disease 2019 (COVID-19) pandemic. The utilization of related services for non-COVID-19 diseases fell dramatically following the point at which the virus broke out; however, little is known about whether this observed decline in healthcare use was due to voluntary behaviors or enforced measures. This paper quantifies the spontaneous change in healthcare utilization during the pandemic. We utilize a county-by-week-level dataset from Taiwan’s National Health Insurance (NHI) record, covering the entire Taiwanese population, and a difference-in-differences design. Our results indicate that even if there were no human mobility restrictions or supply-side constraints, people voluntarily reduced their demand for healthcare, due to fears of contagion, or COVID-related precautionary behaviors. We find that the number of outpatient visits (inpatient admissions) decreased by 21% (11%) during the pandemic period (February to May 2020). Furthermore, the demand response of healthcare for Influenza-like illness (ILI) was much greater and more persistent than for non-ILI, thereby suggesting that the substantial decline in accessing healthcare was induced by positive public health externality of prevention measures for COVID-19. Finally, we find that the demand for healthcare services did not get back to the pre-pandemic baseline, even when there were no local coronavirus cases for 253 consecutive days (mid-April to December 2020) in Taiwan.

## 1 Introduction

COVID-19 has raged through most countries around the world. As of September 2021, it caused 219 million confirmed cases and 4.55 million deaths.^1^ The pandemic has changed many aspects of people’s lives and had negative impacts on macroeconomic activities (Atkeson, 2020; Baker et al., 2020; Eichenbaum, 2020; Guerrieri et al., 2020; Altig et al., 2020), household consumption (Baker et al., 2020; Cicala et al., 2020), and the labor market (Beland et al., 2020; Rojas et al., 2020; Forsythe et al., 2020). Particularly, COVID-19 is a public health crisis, and so healthcare systems have been severely affected during the pandemic. Recent studies have shown that there have been large declines in healthcare utilization for non-COVID-19 diseases (Ziedan et al., 2020; Birkmeyer et al., 2020; Chatterji and Li, 2020; Mehrotra et al., 2020; Hartnett et al., 2020).^2^ However, it is not clear whether this observed decline is due to voluntary behaviors or to other inevitable issues, such as government restrictions on mobility (Chatterji and Li, 2020; Sands et al., 2020; Ziedan et al., 2020) or the availability of health resources (Søreide et al., 2020; Hamel et al., 2020; Rossman et al., 2021).^3^

Measuring the voluntary response in healthcare utilization during the pandemic has important policy implications. First, the pandemic significantly strained the capacity of healthcare systems. To free up medical sources for COVID-19 patients, healthcare providers had to restrict or delay the use of services not related to COVID-19 (Yang et al., 2021). Since such restrictions might also impede the use of some essential medical care and negatively affect people’s health, it raises a question as to what extent the government or healthcare providers have to do so. Previous studies suggest that people change their behaviors voluntarily to reduce the chance of contracting diseases. If the spontaneous response is substantial, the government could achieve a similar outcome by implementing policies with fewer restrictions/costs. Second, compared to policy-induced behavior, voluntary responses could be more persistent. Therefore, understanding voluntary change in healthcare use can help us evaluate the possible impact of disease outbreaks on people’s health behaviors after the pandemic.

This paper fills the gap by examining the effect of the COVID-19 outbreak and pandemic responses on the voluntary demand for non-COVID-19 healthcare. We utilize a difference-indifferences design and a 2014–2020 county-by-week-level dataset from Taiwan’s National Health Insurance (NHI), covering the entire population in Taiwan. Specifically, we examine whether healthcare utilization during the pandemic (2020) varied compared with corresponding weeks in previous years (2014–2019), after controlling for the county-specific trend in demand for healthcare (e.g., county-by-week fixed effects and county-by-year fixed effects).

The case of Taiwan is well-suited for this analysis—for three important reasons. First, as of the end of 2020, Taiwan had only experienced seven deaths and 799 COVID-19 cases, so the pandemic had a very limited impact on healthcare capacity in the country, helping us rule out unmet demand due to supply-side factors. Second, Taiwan did not implement any lockdown or self-isolation policies in 2020; the Taiwanese, for their part, carried on with their normal lives alongside relatively looser regulations. Therefore, our estimated change in demand for healthcare can persuasively represent the voluntary response to the COVID-19 pandemic rather than government restrictions on activity/mobility. Furthermore, Taiwan had a consecutive 253 days of no local cases from April 12^th^ to the end of 2020.^4^ Basically, Taiwanese people returned to normal life during this period. This experience gives us a unique chance to examine whether the behavioral change in healthcare utilization can persist in a “COVID-free” period.^5^ Finally, Taiwan’s NHI is a compulsory single-payer system in which everyone has to enroll; thus, NHI data cover the population-wide healthcare utilization of both outpatient care and inpatient care. This feature allows us to investigate the pandemic effect on different types of healthcare use.

We obtain three key findings from this research. First, our results suggest that on average, the COVID-19 outbreak had larger negative impacts on the utilization of outpatient care (14% decrease) than on inpatient care (4% decrease). In addition, we find that the number of outpatient visits (inpatient admissions) for all diseases decreased by 21% (11%) during the pandemic period (from February to May 2020). Nonetheless, the size of the reduction shrank to 9% for outpatient visits and was null for inpatient admissions in the period when there were no local cases reported (from June to December 2020).

The decline in the voluntary demand for healthcare after the COVID-19 outbreak is likely the combination of two effects. On the one hand, the fear of contracting COVID-19 in hospitals could have led people to reduce or postpone the use of healthcare services that were not urgent or essential. Such an effect should fade away when the virus disappears from the community. On the other hand, COVID-19 prevention measures, such as wearing face mask, might have had an unintended effect by reducing the transmission of Influenza-like illness (ILI) and then decreasing the overall demand for ILI healthcare (i.e., both outpatient and inpatient care). This effect could be large and persistent, since prevention measures for coronavirus indeed mean there is less chance of contracting most ILI diseases, and the Taiwanese still followed these public health measures even when pandemic slowed down.

Second, in order to explore possible mechanisms for our findings, we perform subgroup analyses based on the type of diseases and investigate the dynamic effects of the COVID-19 outbreak. These subgroup analyses suggest that the COVID-19 outbreak led to a much larger decline in healthcare utilization for ILI diseases than for non-ILI diseases (e.g., outpatient visits: 48% vs. 11% decrease, inpatient admissions: 50% vs. 2% decrease). Based on our results, the demand responses of healthcare for non-ILI diseases were likely driven by the fear of catching coronavirus. Because we find that the COVID-19 outbreak had a negative impact on the utilization of outpatient care (i.e., less essential healthcare) but almost no effect on inpatient care (i.e., more essential healthcare). Moreover, COVID-19 effects faded when no new COVID-19 cases were reported.

For ILI diseases, we believe that the large decline in healthcare utilization was mainly caused by the effect of COVID-19 prevention measure, since the demand for both outpatient and inpatient care in terms of ILI diseases experienced similar drops after the COVID-19 outbreak. Furthermore, we find that the negative demand responses persist throughout the whole of 2020. Consistent with the above evidence, our results indicate that the mortality rate of influenza-like illnesses in 2020 was relatively low compared to previous years, thereby suggesting that precautions taken against COVID-19 might have resulted in unintended health benefits.

Third, our estimates imply that voluntary healthcare demand responses induced by the COVID-19 outbreak could “save” on healthcare expenditure by around 52.4 billion NT$ (i.e., 1.9 billion US$), which accounts for 7.3% of the annual healthcare budget.

This paper stands apart from the previous literature in the following ways. First, we contribute to the fast-growing body of literature analyzing the impacts of the COVID-19 pandemic on healthcare systems. Several recent studies indicate that there was a large decline in healthcare utilization during the pandemic period in the US (Ziedan et al., 2020; Birkmeyer et al., 2020; Chatterji and Li, 2020; Mehrotra et al., 2020; Hartnett et al., 2020), UK (Abbas et al., 2021), Spain (Gómez-Ramiro et al., 2021), and Italy (Borrelli et al., 2020).^6^ Our paper provides novel evidence by showing that even if there are no government restrictions on human mobility (e.g., lockdowns or stay-at-home orders) and no supply-side constraints (e.g., inadequate healthcare resources for non-COVID-19 patients), people can voluntarily reduce their demand for healthcare services in response to a virus outbreak. We find that the size of this voluntary response was large during the pandemic (e.g., a 21% decline in outpatient visits) and is comparable to the policy-induced effects on healthcare utilization found in Ziedan et al. (2020), which implies that neglect for voluntary responses may overestimate the effects of policy mandates. Moreover, we provide the first evidence on the persistence of COVID-induced change in health behaviors during the period with no local coronavirus cases.

Second, the spontaneous response to COVID-19 risk broadly relates to the “prevalence response” in the economic epidemiology literature (Ahituv et al., 1996; Gersovitz and Hammer, 2003; Lakdawalla et al., 2006; Paula et al., 2014; Delavande and Kohler, 2012; Bennett et al., 2015).^7^ This paper complements this line of research by showing that people could still take preventive actions proactively to reduce the transmission of the virus, even if prevalence of the disease were low.^8^

Third, this paper is related to the literature on health effects of non-pharmaceutical interventions (NPIs). Most of the recent research points out that such interventions can indeed effectively reduce COVID-19 transmission (Lin and Meissner, 2020; Chernozhukov et al., 2020; Mitze et al., 2020; Abouk and Heydari, 2020; Dave et al., 2020) and might lead to unintended health benefits (Qi et al., 2021; Feng et al., 2021). For example, Feng et al. (2021) found that COVID-19 outbreaks and corresponding NPIs (e.g. lockdown or shelter-in-place order) substantially reduced influenza activity in China and US. However, Taiwan did not implement such high-cost NPIs against COVID-19. Instead, it used relatively low-cost ones, such as wearing face masks and washing hands, and relied on individual distancing efforts. We contribute to this stream of the literature by showing that the interventions based on individual responsibility can still achieve similar health benefits by reducing the incidence of non-COVID-19 diseases such as flu and other pneumonia-based conditions.

## 2 Background

Given its geographic proximity to and close economic relationship with China, where the COVID-19 outbreak began, Taiwan has been considered one of the most affected countries. Surprisingly, upon December 31^st^, 2020, Taiwan had recorded only seven deaths and 799 confirmed cases—671 of which had recovered. The total population of Taiwan is around 23 million so that the country’s infection rate is very low. Moreover, no severe local transmissions had occurred in 2020. Figure 1 displays the number of new confirmed cases for each week of 2020. The first confirmed COVID-19 case was announced in the 4^th^ week of 2020 (i.e., the week of January 21^st^). The dark color represents number of new local confirm cases in a given week and gray color means weekly number of new non-local cases. Fewer than 10% of confirmed cases are local ones, and there have been no new confirmed local cases from the 17^th^ to 51^st^ week of 2020 (i.e., April 12^th^ to December 22^nd^, 2020).

**Figure 1:**
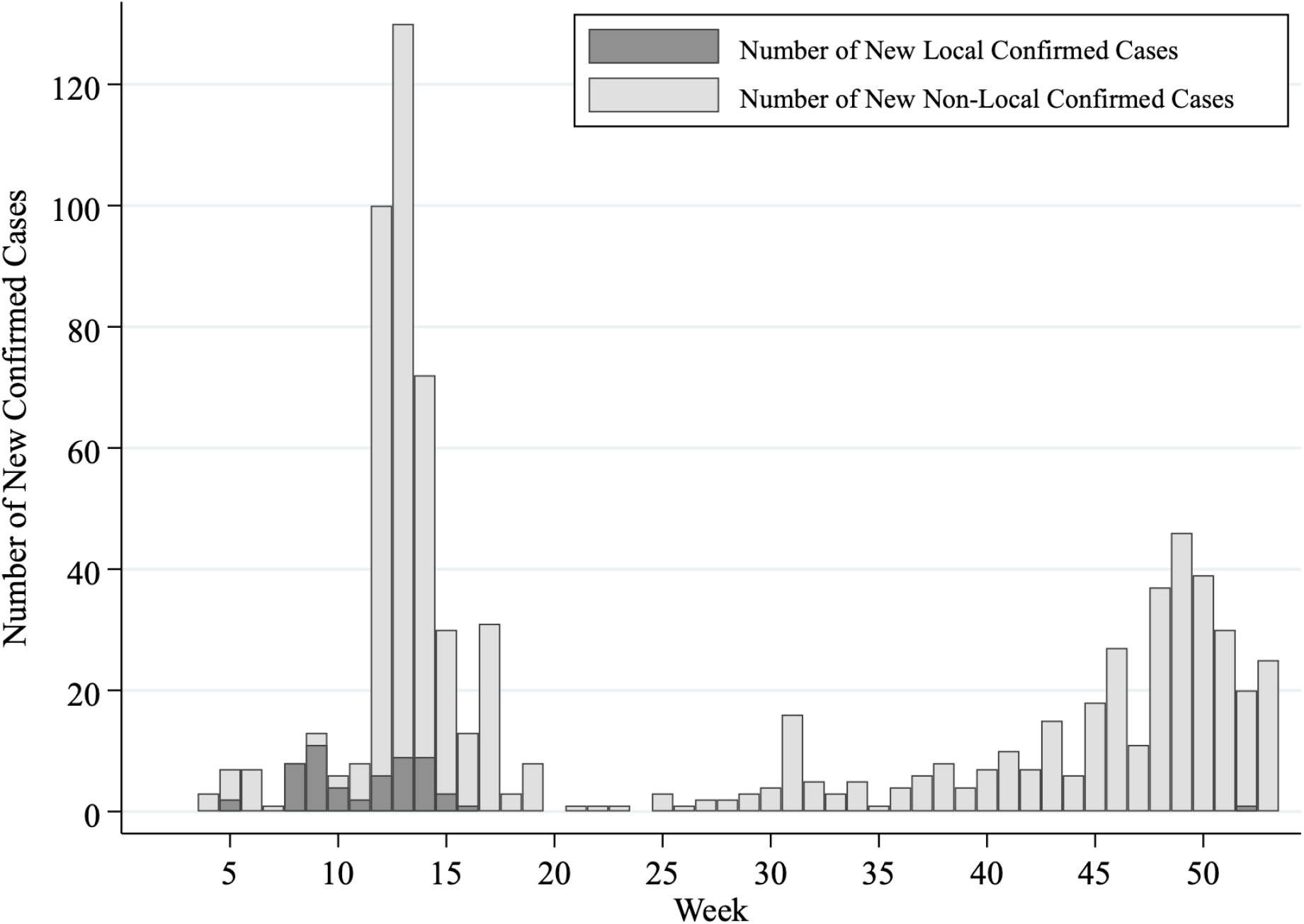
Weekly Number of New Confirmed COVID-19 Cases in Taiwan. *Notes:* This figure displays weekly number of new confirmed COVID-19 cases in Taiwan. The dark color represents number of new local confirm cases in a given week and gray color means weekly number of new imported cases. Data source comes from TCDC.

Basically, COVID-19 pandemic had very limited impacts on people’s life and supply of healthcare services in Taiwan during 2020. On March 25^th^, the government announced a guidance suggesting to cancel unnecessary public gatherings with more than 100 people indoors or 500 people outdoors so that several public events were canceled.^9^ But no lockdown policy, stay-at-home order, or domestic travel ban had been initiated. A few hospitals which were in charge of treating COVID-19 patients did take precautionary measures, such as introducing strict admission and discharge policies, checking patients’ contact and travel history, and delaying (or canceling) non-urgent examinations and procedures. However, given Taiwan’s low incidence rate of COVID-19, the capacities of most healthcare providers were not affected by pandemic. The biggest turning point was on June 7^th^ (i.e., 24^th^ week of 2020). As a consecutive 8 weeks without any local case, the government announced to relax all disease-prevention measures and suggest people can get back to normal life. In general, Taiwan’s government imposed looser regulations on people’s mobility and healthcare system. Instead, it implemented intensive information campaigns to coordinate people’s actions, which were based on individuals’ effort against COVID-19. Using Google Trends data, we find that the Taiwanese people responded to the first reported case by immediately seeking information about the virus and personal protective equipment such as face masks (see Figure A.1 of Online Appendix). Consistent with this finding, a cross-country survey suggests that more than 80% of Taiwanese wore face masks at the beginning of the pandemic (i.e., February, 2020, see Figure A.4 of Online Appendix). These facts suggest that Taiwanese people reacted to COVID-19 in a rapid and proactive way in the very early stages of the outbreak. In the Online Appendix A, we provide more details about how government and residents responded to COVID-19 pandemic.

## 3 Data and Sample

### 3.1 Data

Our healthcare utilization data originate from the Taiwan National ILI Disease Statistics System, accessed via the Taiwan Center for Disease Control’s (TCDC) Open Data Portal.^10^ This database holds NHI claim data, so it covers almost the entire population’s healthcare utilization. In order to investigate the outbreak of selected infectious diseases in a timely manner, the TCDC provides the public with weekly data on the numbers of outpatient visits and inpatient admissions by county, age group, and category of selected infectious diseases. Note that the definition of “week” in this database follows the World Health Organization (WHO)’s definition, which always begins on a Sunday and ends on a Saturday, but does not definitely start from January 1^st^.

To construct our outcome variables—the incidence rate of outpatient visits/inpatient admissions per 100,000 population for specific types of diseases—we divide the number of outpatient visits and inpatient admissions by the population of each corresponding county per year. Population information comes from the population statistics database provided by the Ministry of Interior (MOI), Taiwan.^11^ In our estimated sample, we also include time-varying covariates, such as county-level demographic and weather variables, that may affect health utilization in each county. We acquire demographic data, such as age structure, sex ratio, and educational attainment,^12^ from the MOI statistics portal.^13^ We retrieve daily weather information from the Central Weather Bureau’s (CWB) observation data inquiry system, in order to calculate the weekly average temperature and rainfall for each county.^14^

### 3.2 Sample

The estimated sample is at the weekly-county level. The sample period is from 2014 to 2020, and we use data from the first to the 52^th^ week.^15^ Thus, the estimated sample includes 22 counties *×* 52 weeks *×* 7 years (2014 to 2020), leading to a sample size of 8,008. Furthermore, in order to examine the mechanisms behind COVID-19 effects, we categorize diseases into ILI and non-ILI.^16^

## 4 Empirical Specifications and Results

Our identification strategy is a difference-in-differences (DID) design. Since the first COVID-19 case in Taiwan was reported on January 21^st^ 2020 (i.e., the 4^th^ week of a year), inspired by previous studies (Berniell and Facchini, 2021; Tanaka and Okamoto, 2021; Chang et al., 2020), we use 2020 as the treated year and define the 1^st^ to 3^rd^ weeks and 4^th^ to 52^th^ weeks of the year as the pre-outbreak and post-outbreak periods, respectively. To control for the seasonal pattern of healthcare demand unrelated to the COVID-19 outbreak, we use 2014–2019 as untreated years, in order to construct the counterfactual trend of health utilization in 2020.

Table 1 displays summary statistics for the outcome variables and covariates during the pre-outbreak period (i.e., the first three weeks of a year) and the post-outbreak period (i.e., the 4^th^ to 52^nd^ weeks of a year) in the treated year (i.e., 2020) and untreated years (i.e., 2014–2019). We find that compared to the time trend in untreated years, healthcare utilization experienced a substantial decline after the 4^th^ week in 2020, especially for ILI diseases.

**Table 1:** Summary Statistics for the Treated and Untreated Years

|  | Treated Year<br>2020 |  | Untreated Years<br>2014–2019 |  |
| --- | --- | --- | --- | --- |
|  | Pre-outbreak | Post-outbreak | Pre-outbreak | Post-outbreak |
| **Outcome Variables** (per 100,000 population) |  |  |  |  |
| Number of total outpatient visits | 21,686.64<br>(5,458.21) | 18,713.76<br>(4,904.24) | 20,915.46<br>(5,644.77) | 20,268.92<br>(5,649.82) |
| Number of outpatient visits for ILI diseases | 2,745.49<br>(532.46) | 1,456.45<br>(495.06) | 2,706.18<br>(562.34) | 2,182.31<br>(615.41) |
| Number of outpatient visits for Non-ILI diseases | 18,941.15<br>(5,187.03) | 17,257.31<br>(4,744.46) | 18,209.29<br>(5,360.66) | 18,086.61<br>(5,369.81) |
| Number of total inpatient admissions | 166.86<br>(87.62) | 168.52<br>(88.12) | 148.20<br>(85.92) | 158.38<br>(86.30) |
| Number of inpatient admissions for ILI diseases | 13.48<br>(7.66) | 8.01<br>(4.62) | 10.73<br>(6.90) | 11.29<br>(6.74) |
| Number of inpatient admissions for Non-ILI diseases | 153.38<br>(84.69) | 160.50<br>(85.70) | 137.47<br>(81.37) | 147.09<br>(82.35) |
| Number of ILI deaths | 1.93<br>(0.07) | 1.70<br>(0.03) | 1.54<br>(0.06) | 1.44<br>(0.01) |
| **Demographic Variables** |  |  |  |  |
| Population Size | 1,072,921<br>(1,110,468) | 1,071,819<br>(1,102,253) | 1,068,333<br>(1,091,123) | 1,068,978<br>(1,091,349) |
| Share of Age below 14 | 0.12<br>(0.02) | 0.12<br>(0.02) | 0.13<br>(0.02) | 0.13<br>(0.02) |
| Share of Age between 15 to 64 | 0.72<br>(0.02) | 0.72<br>(0.02) | 0.73<br>(0.02) | 0.73<br>(0.02) |
| Share of Age above 65 | 0.16<br>(0.02) | 0.16<br>(0.02) | 0.14<br>(0.02) | 0.14<br>(0.02) |
| Sex Ratio (Female to Male) | 102.61<br>(8.45) | 102.61<br>(8.45) | 103.25<br>(7.90) | 103.25<br>(7.90) |
| Share of Post-Secondary Degree | 0.44<br>(0.08) | 0.44<br>(0.08) | 0.41<br>(0.08) | 0.41<br>(0.08) |
| Share of High-School Degree | 0.43<br>(0.05) | 0.43<br>(0.05) | 0.44<br>(0.05) | 0.44<br>(0.05) |
| Share of Non-High-School Degree | 0.13<br>(0.04) | 0.13<br>(0.03) | 0.15<br>(0.04) | 0.15<br>(0.04) |
| **Weather Variables** |  |  |  |  |
| Temperature (°C) | 17.12<br>(2.12) | 22.95<br>(4.68) | 16.05<br>(2.69) | 22.65<br>(4.85) |
| Precipitation (mm) | 7.48<br>(10.98) | 4.47<br>(7.13) | 3.34<br>(5.93) | 5.83<br>(9.59) |
| Observations | 66 | 1,078 | 396 | 6,468 |

*Note:* This table displays summary statistics for the outcome variables and covariates during the pre-outbreak period (i.e., the first three weeks of a year) and the post-outbreak period (i.e., the 4<sup>th</sup> to 52<sup>nd</sup> weeks of a year) in the treated year (i.e., 2020) and untreated years (i.e., 2014–2019). Healthcare utilization data comes from the Taiwan National ILI Disease Statistics System, which originates from 2014–2020 NHI claim data. We divide the number of outpatient visits and inpatient admissions by the population of each corresponding county per year to obtain the incidence rate of outpatient visits/inpatient admissions per 100,000 population for specific types of diseases. Demographic information comes from the population statistics database provided by the Ministry of Interior (MOI), Taiwan. Weather variables are from the Central Weather Bureau’s (CWB) observation data inquiry system. Standard deviations are in parentheses.

We first use a conventional DID design to examine the average effects of the COVID-19 outbreak on healthcare utilization, following which we extend the design to a multiple period DID and an event study design, to investigate further the dynamic trajectory of COVID-19 effects.

### 4.1 Graphical Evidence

Figure 2 displays trends in the utilization of outpatient care and inpatient care. The solid line represents the trend in 2020, and the dashed line denotes the average across 2014–2019 with a 95% confidence interval. Since the first confirmed COVID-19 case was announced in the 4^th^ week of 2020, the vertical line in the graph separates that week from the following weeks. The vertical axis in Figure 2 stands for the percentage change in the number of outpatient visits (inpatient admissions) from the baseline week, which is the second week of each year.

**Figure 2:**
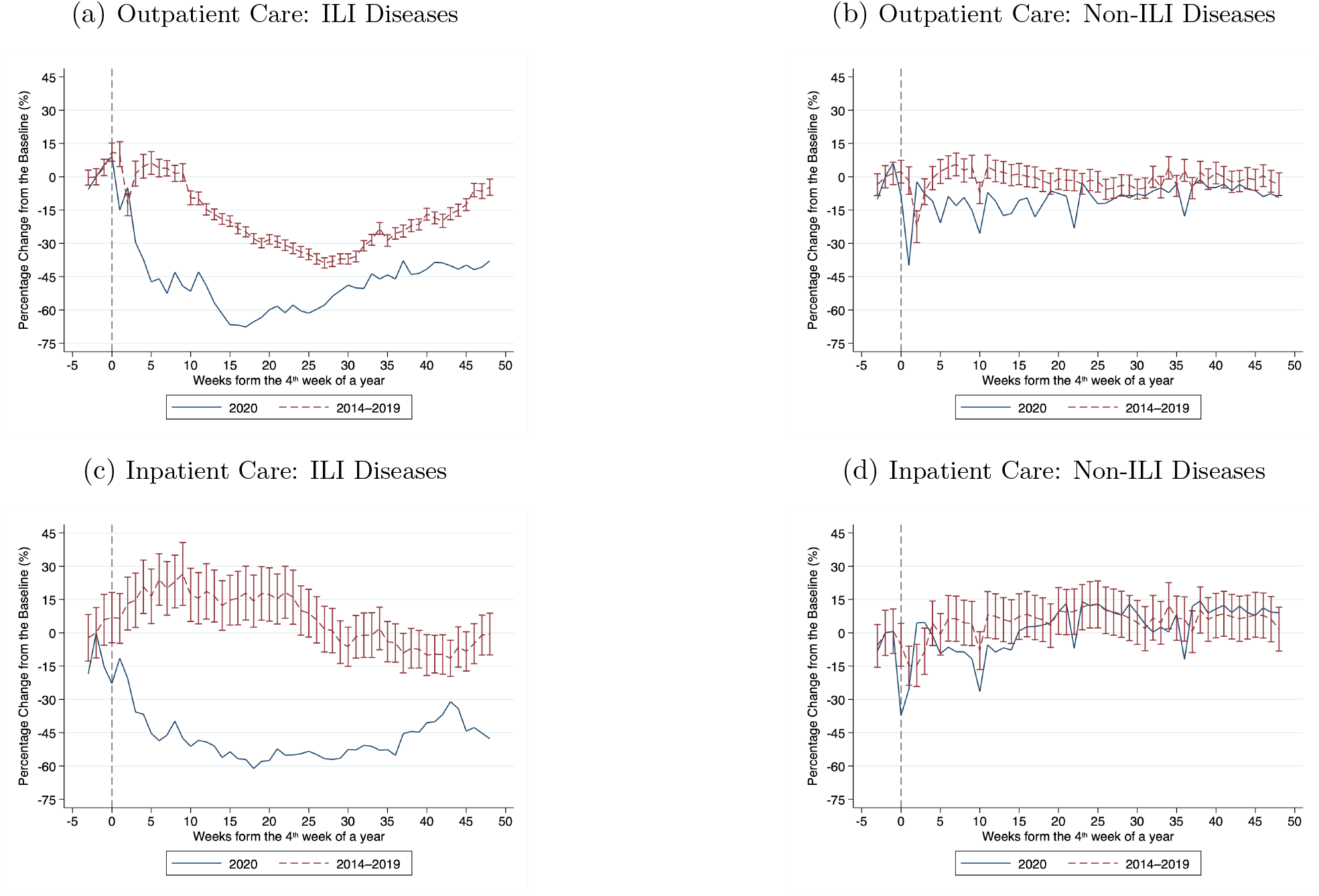
Percentage Change in Number of Outpatient Visits/Inpatient Admissions. *Notes:* Sample period is 2014–2020. The solid line represents the trend in 2020, and the dashed line denotes the average across 2014–2019 with a 95% confidence interval (error bar). Since the first confirmed COVID-19 case was announced in the 4^*th*^ week of 2020 (i.e. the week of January 21^*st*^), the vertical line in the graph separates that week from the following weeks. The vertical axis of this figure stands for the percentage change in the number of outpatient visits (inpatient admissions) from the baseline weeks. We use the number of outpatient visits (inpatient admissions) in the second week of each year as the baseline.

Figure 2a shows the trend in outpatient visits for ILI diseases. As the flu season usually runs from October to April in Taiwan, numbers of visits for ILI diseases tend to be higher until April during 2014–2019 (see the dashed line). In a sharp deviation from the usual seasonal patterns of 2014–2019, the numbers of ILI visits fell by 30% to 65% after the COVID-19 outbreak, and the declining pattern was still persistent at the end of 2020 (see the solid line). For non-ILI diseases, Figure 2b shows that there was a large decline in visits for non-ILI diseases during the lunar new year (i.e., the 5^*th*^ week of 2020) and a rebound after the holiday. This pattern can also be found in 2014–2019. The difference between 2020 and the previous few years is that the average numbers of visits for non-ILI diseases in 2020 did not rebound back to the baseline level, and in fact they declined by around 15% until the middle of the year.

Figure 2c illustrates the evolution of inpatient utilization for ILI diseases. Compared to the baseline weeks, admissions for ILI diseases decreased by 15% to 60% after the virus outbreak in 2020, which is very different from the trends in 2014–2019. In contrast, Figure 2d suggests inpatient admissions for non-ILI diseases in 2020 largely followed patterns similar to those in 2014–2019 except that there was a small decline during the 6^th^ week to the 13^*th*^ week after the first COVID-19 case.

### 4.2 COVID-19 Effects on Healthcare Utilization

In order to obtain average effect of COVID-19 outbreak on healthcare utilization, we estimate the following DID specification:

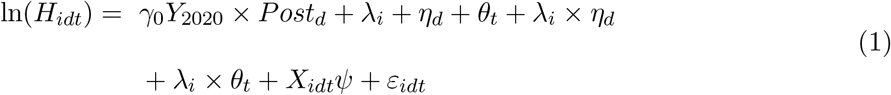

Our estimation is implemented at the weekly-county level. *H*_*idt*_ represents the outcomes of interest, namely, numbers of outpatient visits (inpatient admissions) per 100,000 population in county *i* in week *d* of year *t*. We focus on three measures of healthcare utilization: 1) visits/admissions for all diseases; 2) visits/admissions for ILI diseases; and 3) visits/admissions for non-ILI diseases. *Y*_2020_ is a dummy for the treated year that takes one if the observation is in 2020, and zero otherwise. *Post*_*d*_ is a dummy indicating the post-outbreak period (after the 4^th^ week of a year). We include the county fixed effect *λ*_*i*_ to control for any time-invariant confounding factors at the county level. The week-of-the-year fixed effect *η*_*d*_ controls for seasonal patterns in healthcare utilization at the national level within a year and year fixed effect *θ*_*t*_ controls for the general trend in healthcare utilization over time. To account for any county-specific seasonal patterns or health shocks, we include the county-by-week fixed effect (*λ*_*i*_ *× η*_*d*_) and the county-by-year fixed effect (*λ*_*i*_ *× θ*_*t*_). *X*_*idt*_ refers to a set of covariates, including various holiday dummies (e.g., the Lunar New Year week) and county-level variables, such as age structure, sex ratio, educational attainment, average temperature, average rainfall, and county-specific linear time trend.^17^ We estimate the equation (1) using a Poisson regression since the outcome variable is weekly count of outpatient visits/inpatient admissions. In order to account for possible within-group correlations of errors, we use the multiway clustering approach proposed by Cameron et al. (2012) to calculate standard errors clustered at both the year-week and the county levels. Finally, all regressions are weighted by the monthly population size of a county. In Section 4.5, we conduct robustness checks on the estimates by utilizing different specifications and computing the standard errors at different cluster levels.

The key variable used for identification in the equation (1) is an interaction term between an indicator for the treated year *Y*_2020_ and a dummy for the post-outbreak period *Post*_*d*_. The coefficients of interest are *γ*_0_, measuring the difference in healthcare utilization before and after the COVID-19 outbreak in 2020 (i.e., the treated year), relative to the difference in the corresponding periods for 2014–2019 (i.e., the untreated years). *γ*_0_ can represent COVID-19 effects on healthcare utilization if the common trend assumption holds. That is, in the absence of the COVID-19 outbreak, the weekly trend in healthcare utilization should be similar in the treated and the untreated years. We examine this assumption by using the DID event study design and a set of placebo tests.

Table 2 shows the DID estimates (i.e., the coefficient on *Y*_2020_ *× Post* of equation (1)). The first four columns display our results for outpatient care. We gradually include control variables to test the sensitivity of the results to different specifications. Estimates across the specifications are fairly independent of the introduction of different sets of covariates and fixed effects. Our preferred specification is in Column (4) of Table 2, which includes a full set of covariates.^18^ The estimate in Column (4) of Panel A suggests that compared to the same weeks in 2014–2019, total outpatient visits during the post-outbreak period in 2020 significantly decreased by 14%. Furthermore, the estimates in Column (4) of Panels B and C show that ILI visits saw a much larger decline (i.e., 48% decrease) than non-ILI visits (i.e., 11% decrease).

**Table 2:** Effects of COVID-19 Outbreak on Non-COVID-19 Health Utilization

|  | (1) | (2) | (3) | (4) | (5) | (6) | (7) | (8) |
| --- | --- | --- | --- | --- | --- | --- | --- | --- |
|  | Outpatient Care |  |  |  | Inpatient Care |  |  |  |
| **Panel A:** All diseases |  |  |  |  |  |  |  |  |
| $Y_{2020} \times Post$ | -0.14***<br>(0.01) | -0.13***<br>(0.02) | -0.14***<br>(0.02) | -0.14***<br>(0.02) | -0.05***<br>(0.00) | -0.04***<br>(0.01) | -0.05<br>(0.03) | -0.04<br>(0.03) |
| **Panel B:** ILI diseases |  |  |  |  |  |  |  |  |
| $Y_{2020} \times Post$ | -0.46***<br>(0.04) | -0.47***<br>(0.04) | -0.47***<br>(0.04) | -0.48***<br>(0.04) | -0.51***<br>(0.02) | -0.52***<br>(0.05) | -0.51***<br>(0.07) | -0.50***<br>(0.06) |
| **Panel C:** Non-ILI diseases |  |  |  |  |  |  |  |  |
| $Y_{2020} \times Post$ | -0.10***<br>(0.00) | -0.10***<br>(0.02) | -0.11***<br>(0.02) | -0.11***<br>(0.02) | -0.02***<br>(0.00) | -0.01<br>(0.02) | -0.02<br>(0.03) | -0.02<br>(0.03) |
| Observation | 8,008 |  |  |  |  |  |  |  |
| Basic control | ✓ | ✓ | ✓ | ✓ | ✓ | ✓ | ✓ | ✓ |
| Demographic variables |  | ✓ | ✓ |  |  | ✓ | ✓ |  |
| Weather variables |  | ✓ | ✓ | ✓ |  | ✓ | ✓ | ✓ |
| County fixed effect |  |  | ✓ | ✓ |  |  | ✓ | ✓ |
| County-by-year fixed effect |  |  |  | ✓ |  |  |  | ✓ |
| County-by-week fixed effect |  |  |  | ✓ |  |  |  | ✓ |
| County specific time trend |  |  |  | ✓ |  |  |  | ✓ |

Columns (5) to (8) display the DID estimates for inpatient care. Column (8) of Table 2 provides our preferred estimates. The estimate in Column (8) of Panel A indicates that the COVID-19 outbreak reduced total inpatient admissions by 4%. This estimate is only one-third of the decline in outpatient care. The estimates in Column (8) of Panels B and C show the results for ILI admissions and non-ILI admissions, respectively. Compared to the trend in previous years, the number of ILI admissions declined by 50% during the pandemic period in 2020 (see Panel B). In contrast, there was only a negligible decrease in inpatient admissions for non-ILI diseases (see Panel C).

### 4.3 COVID-19 Effects During and After the Pandemic

In 2020, Taiwan had no local COVID-19 cases from April 12^th^ to the end of 2020 (i.e., around 253 days), so the government relaxed preventive measures for COVID-19 on June 7^th^ (i.e., the 24^th^ week of 2020) and suggested that people could return to a normal life. Therefore, we divide the post-outbreak period into 1) pandemic period and 2) COVID-free period, and estimate the following multi-period DID specification:

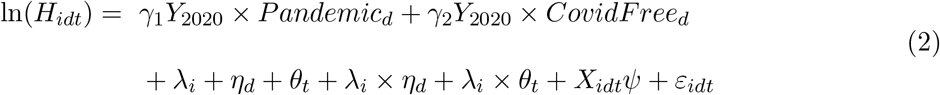

*Pandemic*_*d*_ denotes the dummy variable, which takes a value of 1 during the 4^th^ to the 23^rd^ weeks (the pandemic period). *CovidFree*_*d*_ takes the value 1 during 24^th^ to the 52^nd^ weeks (COVID-free period). The coefficients of interest are *γ*_1_ (*γ*_2_), which measure changes in healthcare utilization during the pandemic period (COVID-free period) compared to the pre-outbreak period (the first three weeks of a year) in 2020, relative to the corresponding weeks in 2014-2019. If the COVID-19 effects faded out when there were no local cases, the *γ*_2_ should shrink to zero.

The first four columns of Table 3 illustrate the results for outpatient care. Again, we gradually include covariates to examine the robustness of our estimates to various specifications. The DID estimates in all specifications are fairly stable. The estimate in Column (4) of Panel A suggests that compared to the same weeks in 2014–2019, total outpatient visits during the pandemic period in 2020 significantly decreased by 21%. In addition, we find that the reduction in outpatient utilization persisted during the COVID-free period, but the estimate shrank to a 9% decline. The estimates in Panel B indicate that ILI visits experienced a large decline in both the pandemic period (57% decrease) and the COVID-free period (40% decrease). However, we find that outpatient visits for non-ILI diseases saw a 17% drop during the pandemic period and rebounded to a 7% reduction during the COVID-free period (see Panel C).

**Table 3:**
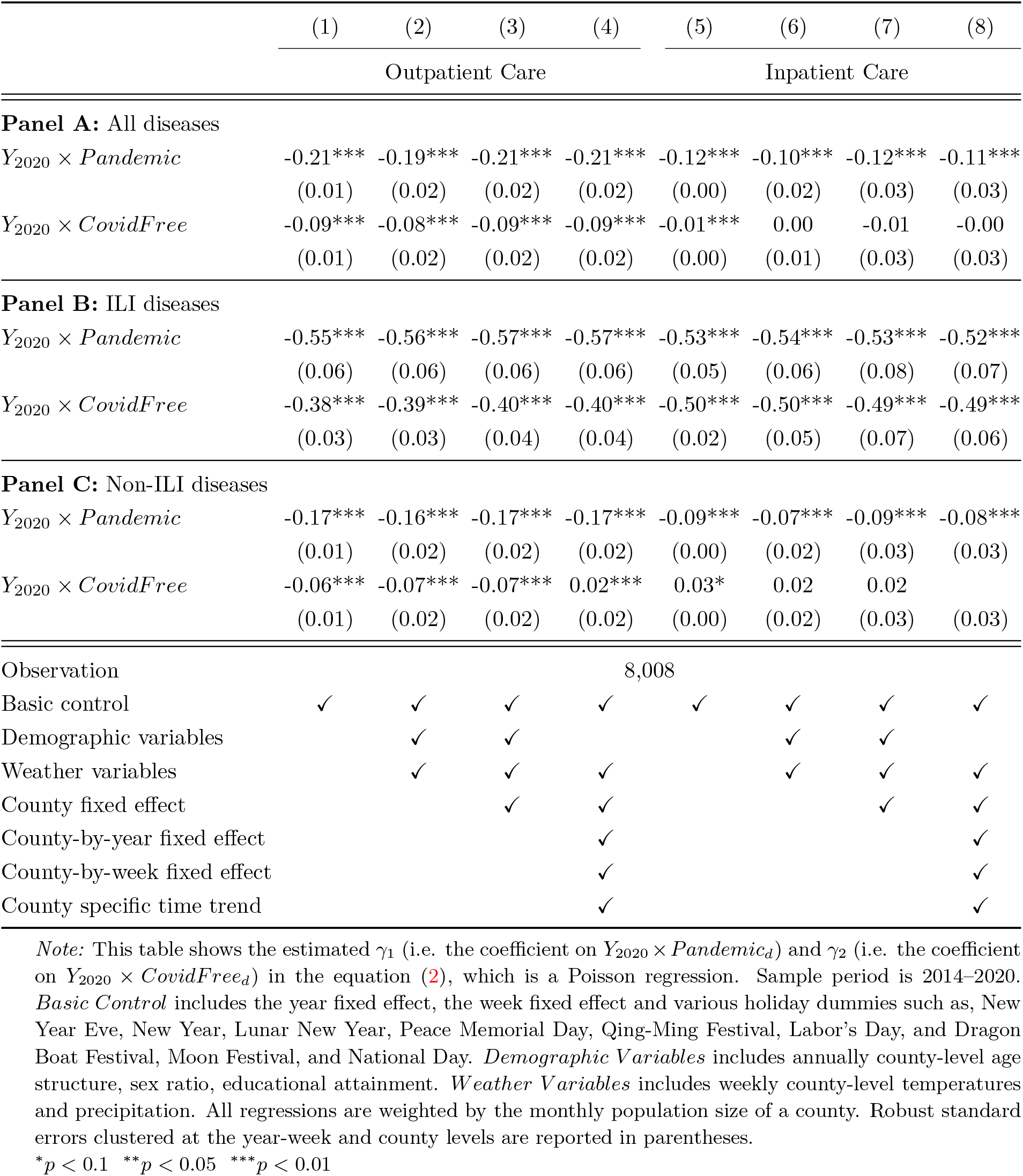
Effects of COVID-19 Outbreak on Health Utilization (by Pandemic Periods)

The last four columns of Table 3 show the results for inpatient care. Column (8) of Table 3 is our preferred specification. Panel A indicates that the COVID-19 outbreak did indeed significantly reduce total inpatient admissions by 11% in the pandemic period. In contrast, the COVID-19 impact disappeared during the period when Taiwan had no local COVID-19 cases (point estimate is zero and statistically insignificant). Interestingly, the estimates in Panel B suggest that the decline in inpatient utilization for ILI diseases continued during both the pandemic period (52% decrease) and the COVID-free period (49% decrease). Similar to the results in Panel A, inpatient admissions for non-ILI diseased only declined during the pandemic period (8% decline) but rebounded to pre-pandemic level in the period without local virus cases.

### 4.4 Dynamic Effects of COVID-19 on Healthcare Utilization

In order to examine common trend assumption and outline the full dynamic trajectory of the COVID-19 effects, we implement an event study design by interacting the treated year dummy *Y*_2020_ with lead and lag time dummies *W*_*d*_.

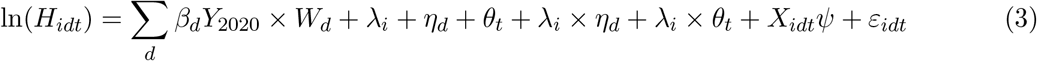

We use *W*_*d*_, where *d* = *−*3, *−*1, 0, 1, 2, 3, ….25, 26, 48, to denote dummy variables for the weeks before and after the 4^th^ week of a year. For example, *W*_1_ represents a dummy for the first week after the announcement of the first confirmed COVID-19 case. Note that we use the 2^nd^ week of a year as the baseline week (i.e., *d* = *−*2).

The key variables used for identification in equation (3) are a set of week dummies *W*_*d*_ interacted with the treated year dummy *Y*_2020_. The coefficients of interest are *β*_*d*_, measuring the difference in healthcare utilization between week *d* and the baseline week for 2020 (i.e., the treated year), relative to the difference for 2014–2019 (i.e., the untreated years). *β*_*d*_ can represent the dynamic effect of the COVID-19 outbreak on healthcare utilization. Figure 3 highlights the estimated *β*_*d*_ in equation (3), which measures the dynamic effect of the COVID-19 outbreak on healthcare utilization, and the corresponding 95% confidence intervals. The horizontal axis denotes the number of weeks from the COVID-19 outbreak (i.e., the 4^th^ week in a year). The top (bottom) panel of Figure 3 displays the results for outpatient (inpatient) care.

**Figure 3:**
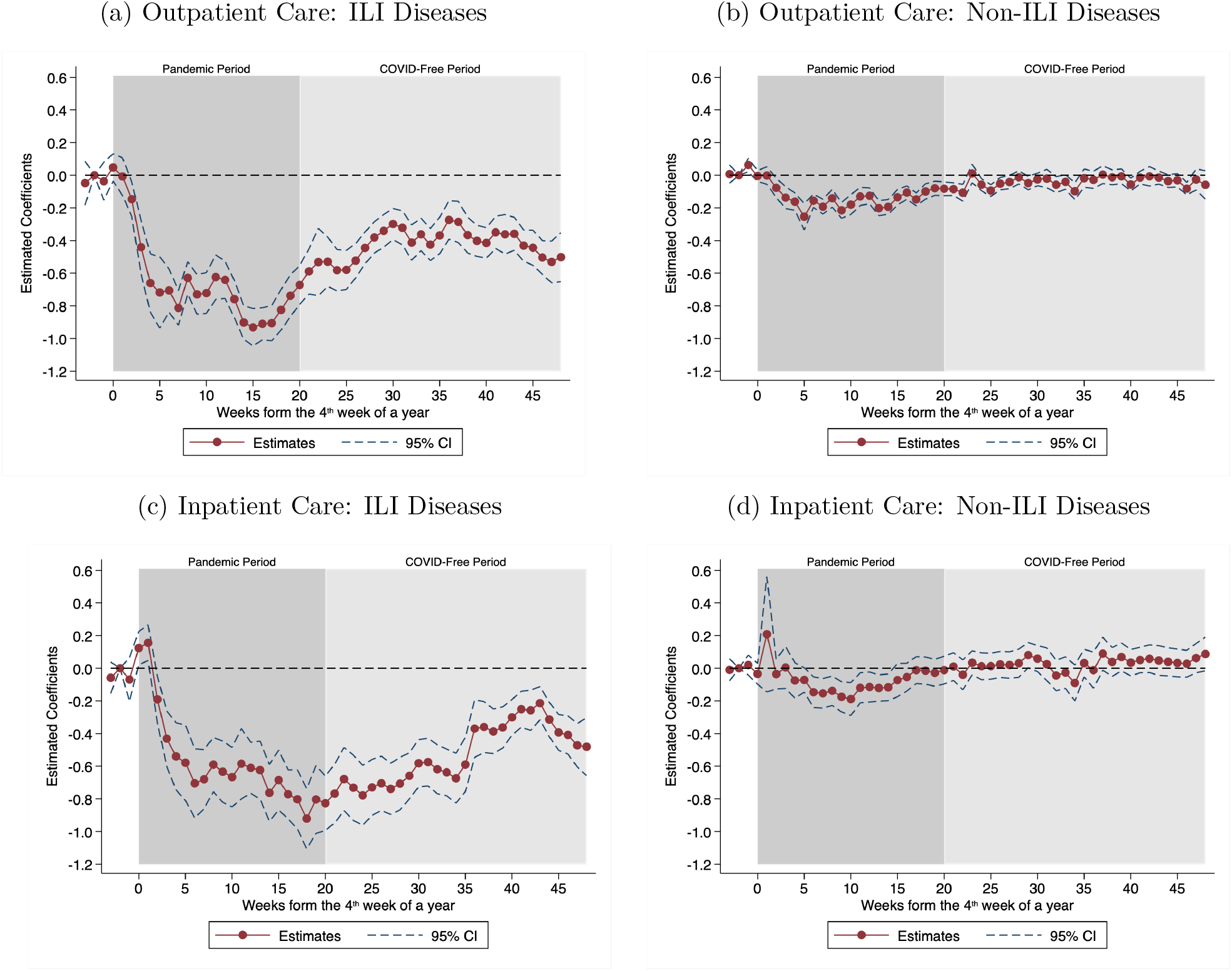
Dynamic Effects of COVID-19 Outbreak on Non-COVID-19 Health Utilization. *Notes:* Figure 3 shows the estimated *β*_*d*_ in equation (3). The dashed lines represent the corresponding 95% confidence intervals. The horizontal axis denotes the number of weeks from the COVID-19 outbreak (i.e., the 4^th^ week in a year). The top (bottom) panel of Figure 3 displays the results for outpatient (inpatient) care by disease type. In order to account for possible within-group correlations of errors, we use the multiway clustering approach proposed by Cameron et al. (2012) to calculate standard errors clustered at both the year-week and the county levels. All regressions include county-by-year fixed effects and county-by-week fixed effects and are weighted by the monthly population size of a county. Sample size is 8,008 and sample period is 2014–2020.

Three key insights emerge from the figures. First, estimates for the first three weeks of a year (i.e., *d* = *−*3, *−*2, *−*1) in all figures are close to zero, suggesting that trends in the numbers of outpatient visits/inpatient admissions between the treated year (i.e., 2020) and the untreated years (i.e., 2014–2019) were in parallel before the COVID-19 outbreak. Therefore, the common trend assumption of our DID design is valid.

Second, Figure 3a indicates that the size of the reduction in visits for ILI diseases is very large. The COVID-19 outbreak reduced the utilization of outpatient care for ILI diseases by about 70% within the first four weeks of the pandemic, and these effects then persisted, thereby suggesting that ILI disease visits still declined by at least 50% at the end of the sample period. For visits in relation to non-ILI diseases, Figure 3b suggests that the reduction in outpatient use is relatively smaller. The number of visits for non-ILI diseases declined by 20% in the 4^th^ week after the first case was reported and shrank to zero in late June (i.e., the 23^*rd*^ week after first COVID-19 case), because there were no local COVID-19 cases in Taiwan for around two months.

Third, Figure 3c suggests that the number of inpatient admissions for ILI diseases decreased by about 50% in the 4^th^ week after the announcement of the first COVID-19 case. Consistent with outpatient care, the reduction in ILI diseases admissions never rebounded to the pre-pandemic level until the end of 2020. Figure 3d indicates that inpatient admissions for non-ILI diseases significantly declined by around 20% when the number of COVID-19 cases accumulated quickly and reached peak (i.e., the 6^th^ week to the 13^*th*^ week after the first COVID-19 case). Interestingly, the COVID-19-induced decline in the number of admissions for non-ILI diseases dropped immediately to zero when Taiwan began to have no local case (i.e., the 14^*th*^ week after the first COVID-19 case).

### 4.5 Placebo Test and Robustness Checks

In this section, we first implement a series of placebo tests by excluding observations in 2020 and only using the 2014–2019 sample. Following previous studies (Tanaka and Okamoto, 2021; Leslie and Wilson, 2020; Heft-Neal et al., 2020), we randomly select one year as the pseudo “treated year” for each county and estimate equation (1). We repeat the above procedures 1,000 times to obtain the distribution of placebo estimates. Figure 4 compares the real estimate with these placebo estimates. Our results suggest that for the outpatient care of all diseases and inpatient care of ILI diseases, the real estimates are way below the placebo ones (see Figure 4a, 4c, 4d, and 4e). In sum, this placebo test indicates that the significant estimates in Table 2 should be treated as causal and are not just findings made by chance.

**Figure 4:**
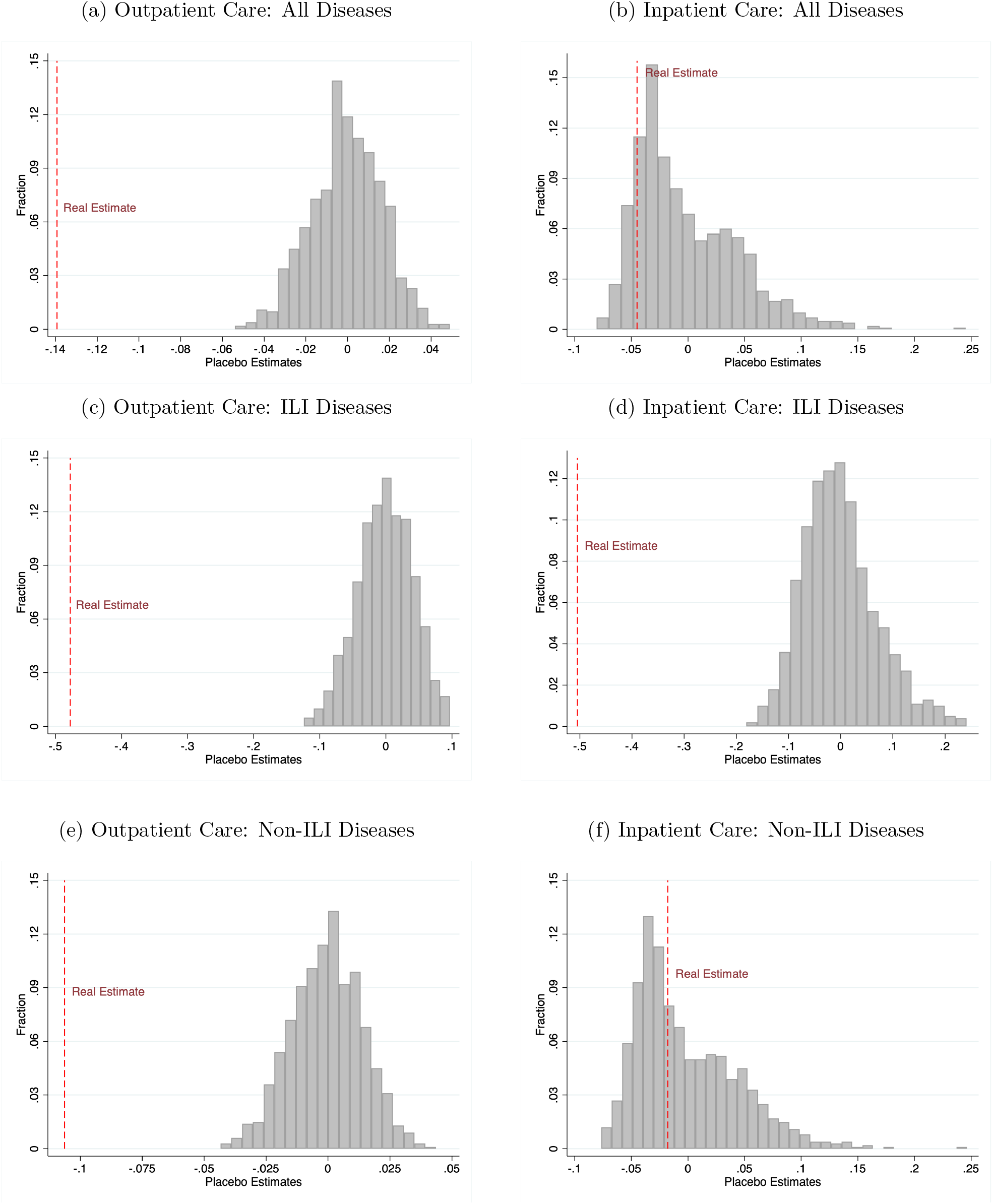
Placebo Test: DID Design. *Notes:* We use the 2014–2019 sample and randomly select one year as the pseudo “treated year” in each county and estimate equation (1). We repeat the above procedures 1,000 times to obtain the distribution of placebo estimates. Figure 4 compares our real estimate with these placebo estimates. The vertical axis displays the relative frequency of the estimates. The gray bars denote the placebo estimates, and the red dashed lines denote the real ones.

We perform the same placebo tests for our multi-period DID design (equation (2)) and event-study analysis (equation (3)). Figure 5 displays the results for equation (2). The red and blue dashed lines denote the real estimates for COVID-19 effects during the pandemic period and COVID-free period, respectively. The placebo test verifies that all significant estimates in Table 3 are not the result of randomness. Figure 6 illustrates the results for equation (3). The red lines show real estimates, and the gray lines denote 1,000 placebo ones. Again, the falsification test confirms that significant estimates in the event-study analysis are unlikely to be chance findings.

**Figure 5:**
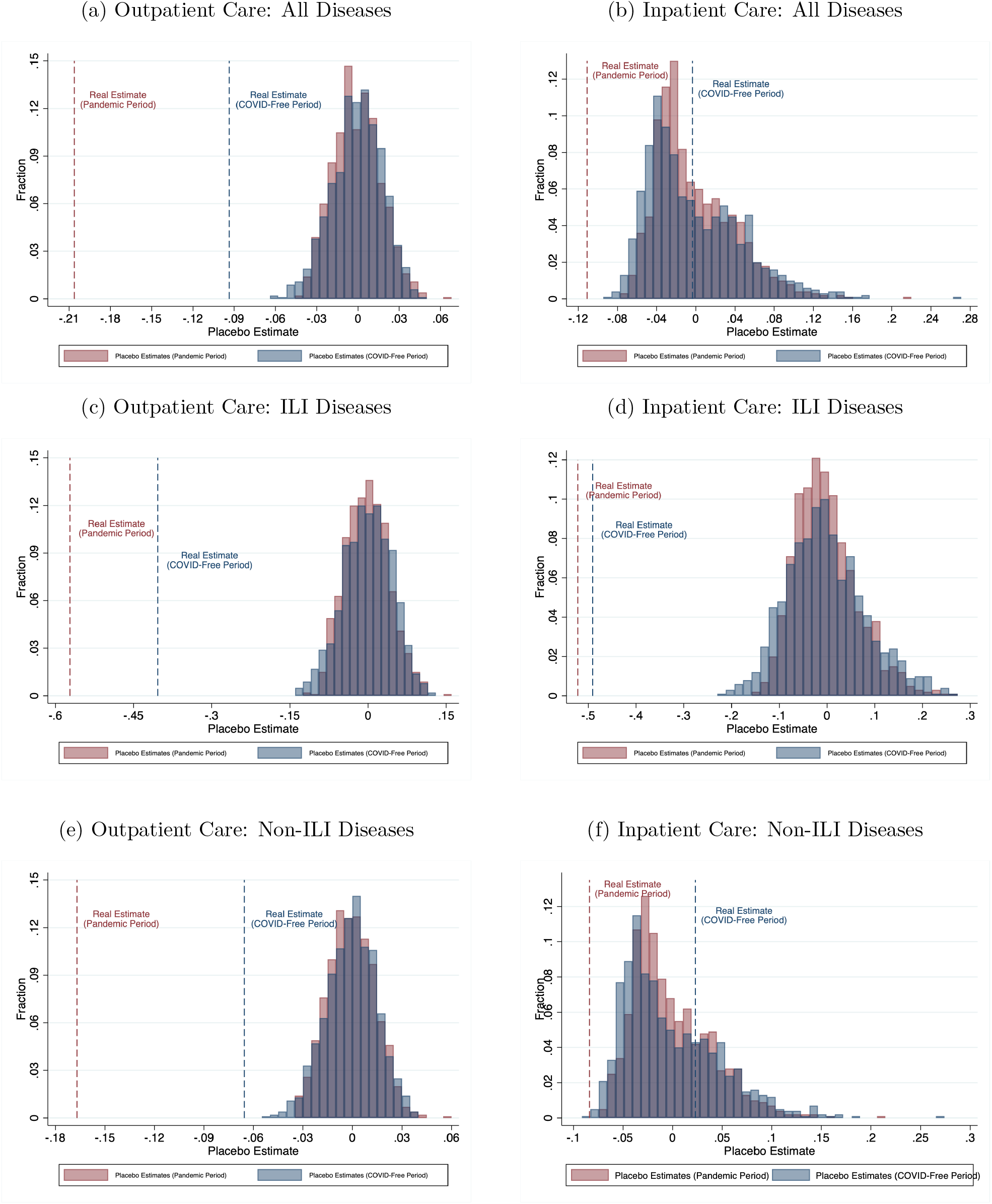
Placebo Test: Multi-Period DID Design. *Notes:* We use the 2014–2019 sample and randomly select one year as the pseudo “treated year” in each county and estimate equation (2). We repeat the above procedures 1,000 times to obtain the distribution of placebo estimates. Figure 5 compares our real estimate with these placebo estimates. The vertical axis displays the relative frequency of the estimates. The red (blue) dashed line denotes the real estimates for COVID-19 effects during the pandemic period (COVID-free period). The red (blue) bars denote the placebo estimates for COVID-19 effects during the pandemic period (COVID-free period).

**Figure 6:**
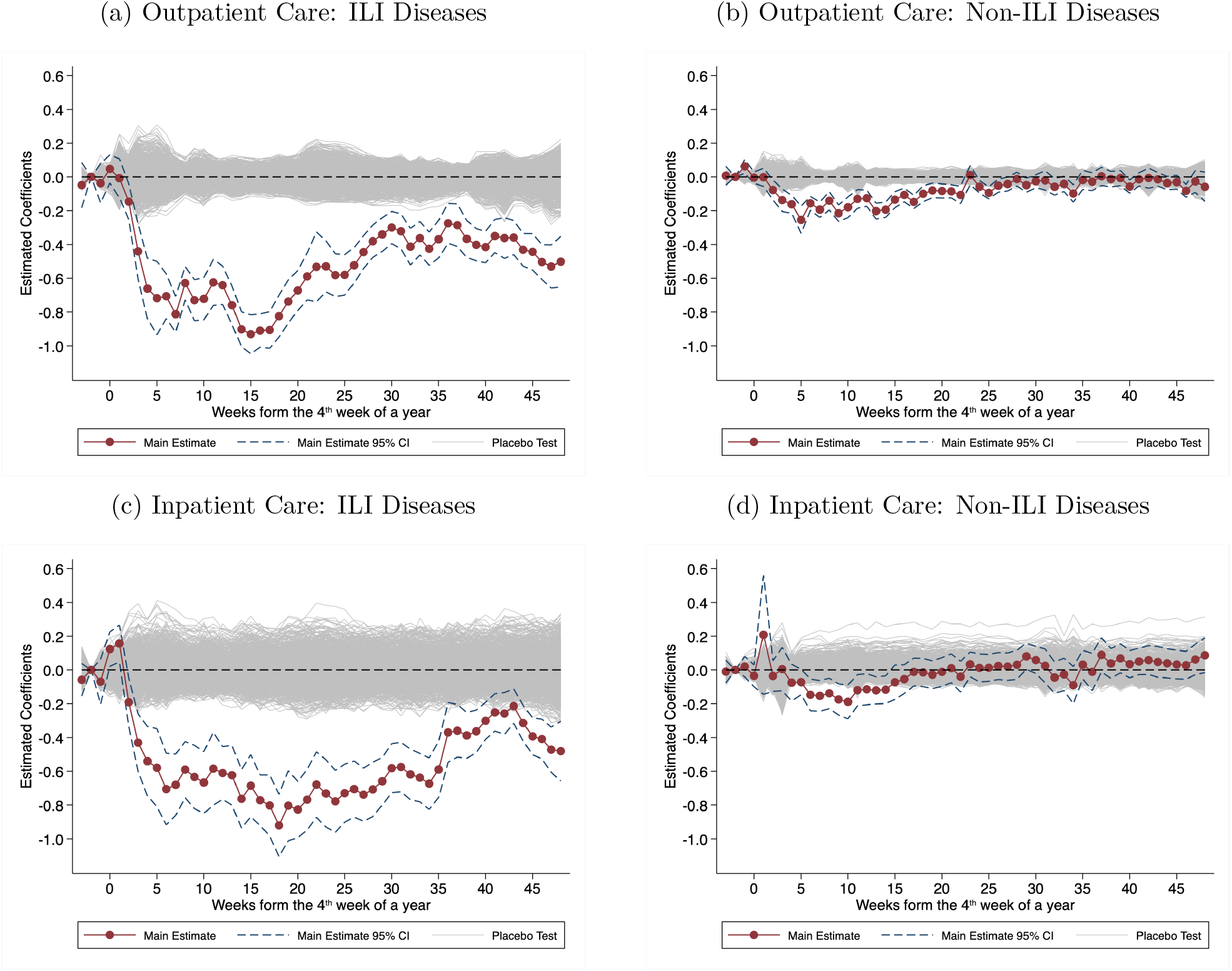
Placebo Test: Event-study Design. *Notes:* We use the 2014–2019 sample and randomly select one year as the pseudo “treated year” in each county and estimate equation (3). We repeat the above procedures 1,000 times to obtain the distribution of placebo estimates. Figure 6 compares our real estimates with these placebo estimates. The gray lines denote the placebo estimates, and the red dots and blue dashed lines denote the real estimates and the corresponding 95% confidence intervals.

Next, in the Online Appendix B, we conduct several robustness checks, using various specifications. First, we calculate standard errors based on different clustering levels, in order to examine the robustness of statistical inference. In our main specification, we cluster the standard error at both the year-week and the county levels. We also conduct statistical hypothesis tests using standard errors clustered on the county or year-week levels, respectively (see Table B.1 and Table B.2). We find that the statistical significance of the estimates is robust to the standard errors clustered at different levels. Second, Table B.3 and B.4 show the estimates based on unweighted Poisson regression (equation (1) and (2)). Our main results are robust to this change.

## 5 Discussion

### 5.1 Interpretation of the Results

So far, we have found that the COVID-19 outbreak was associated with a substantial reduction in both outpatient and inpatient utilization. In addition, the healthcare utilization for ILI diseases experienced a much larger decline than for non-ILI diseases. Further, negative impacts peaked during the pandemic period, and then began to rebound when there were no local cases. Since COVID-19 had limited impacts on Taiwan’s healthcare system, and the government did not implement any mobility-restricted policy or close health facilities, the reduction in healthcare utilization is unlikely to have been caused by issues related to human mobility restrictions or healthcare supply.

Therefore, the decline in demand for healthcare during the pandemic period is likely to be voluntary responses, which are mixed with two effects, namely the fear effect and the prevention effect. First, people may have reduced healthcare utilization due to the fear of COVID-19 infection. As hospitals are usually a place with a high risk of contracting diseases, patients might postpone or cancel their visits if receiving medical treatment is neither necessary nor urgent: we call this mechanism the “fear effect.” Second, since ILI diseases (e.g., flu or other forms of pneumonia) shares many similarities with COVID-19 in terms of disease presentation and the ways of transmission, COVID-19 prevention measures, such as wearing a face mask, hand-washing, and social distancing, might have had an unintended effect by reducing the transmission of ILI diseases. Thus, the demand for healthcare could have decreased due to an improvement in health status. This particular mechanism is called the “prevention effect.”

In our main analysis, we find that the COVID-19 outbreak caused different impacts on healthcare utilization for ILI diseases and non-ILI diseases. Furthermore, the negative impact of the COVID-19 outbreak was most severe during the pandemic period and faded out when there was very little risk of the local spread of COVID-19 in Taiwan. Such differences help us understand the mechanisms behind healthcare demand responses to the COVID-19 outbreak. If the decline in healthcare utilization were mainly driven by the fear effect, we should expect that the COVID-19 outbreak would have had less of a negative impact on the utilization of inpatient care (i.e., more essential healthcare) than of outpatient care (i.e., less essential healthcare). In addition, the demand response to the fear of contracting COVID-19 might have disappeared when the risk of catching the virus was low.

Our results indicate that the demand response of healthcare for non-ILI diseases could have been induced by the fear effect of contracting COVID-19. For non-ILI diseases, the COVID-19 outbreak led to a 11% reduction in outpatient visits but almost no impact on inpatient admissions (i.e., insignificant 2% decrease). Furthermore, we find that the negative effect of the COVID-19 outbreak on the utilization of both outpatient care and inpatient care for non-ILI diseases faded out when no new local COVID-19 cases were reported in Taiwan. Interestingly, the event study analysis indicates that the demand response of inpatient care vanished earlier than for outpatient care (i.e., the 14^*th*^ week vs. the 23^*rd*^ week after the first COVID-19 case).

On the other hand, if the reduction in healthcare demand was mainly driven by the effect of COIVD-19 preventive measures, we should expect that the utilization of both outpatient and inpatient care declined similarly because these measures basically helped stop the spread of ILI diseases. In addition, the negative effect should be sizable and persistent since Taiwanese people maintain these healthy habits during the year 2020. For example, Figure A.4 of the Online Appendix A shows that the proportion of people who wear a face mask in public spaces remained at over 80% throughout the whole of 2020.

We do find that there were large declines in both outpatient visits and inpatient admissions for ILI diseases after COVID-19 outbreak and the negative effects continued even during the period when Taiwan had no local COVID-19 case. Moreover, if precautions against coronavirus indeed led to a decline in ILI-related healthcare use, we should also find corresponding evidence on improvements in health status, such as a decline in ILI mortality.

Figure 7 displays the percentage change in the weekly number of ILI-related deaths per 100,000 population from the baseline mean (i.e., the average outcome of the second weeks in each year). The solid line represents the trend of ILI-related mortality rate in 2020. The dashed line represents the 12-year average of the outcome during 2008–2019 and the corresponding 95% confidence interval. Again, the vertical line in the graph denotes the 4^th^ week of a year. Our results suggest that the trend for the ILI-related mortality rate in 2020 is an outlier compared to the same period in the previous 12 years. The percentage change from the baseline mean for the ILI-related mortality rate fell by 10% to 30% after COVID-19 outbreak. This result is consistent with the finding in recent literature on unintended health benefits of NPIs against COVID-19 (Qi et al., 2021; Feng et al., 2021).

**Figure 7:**
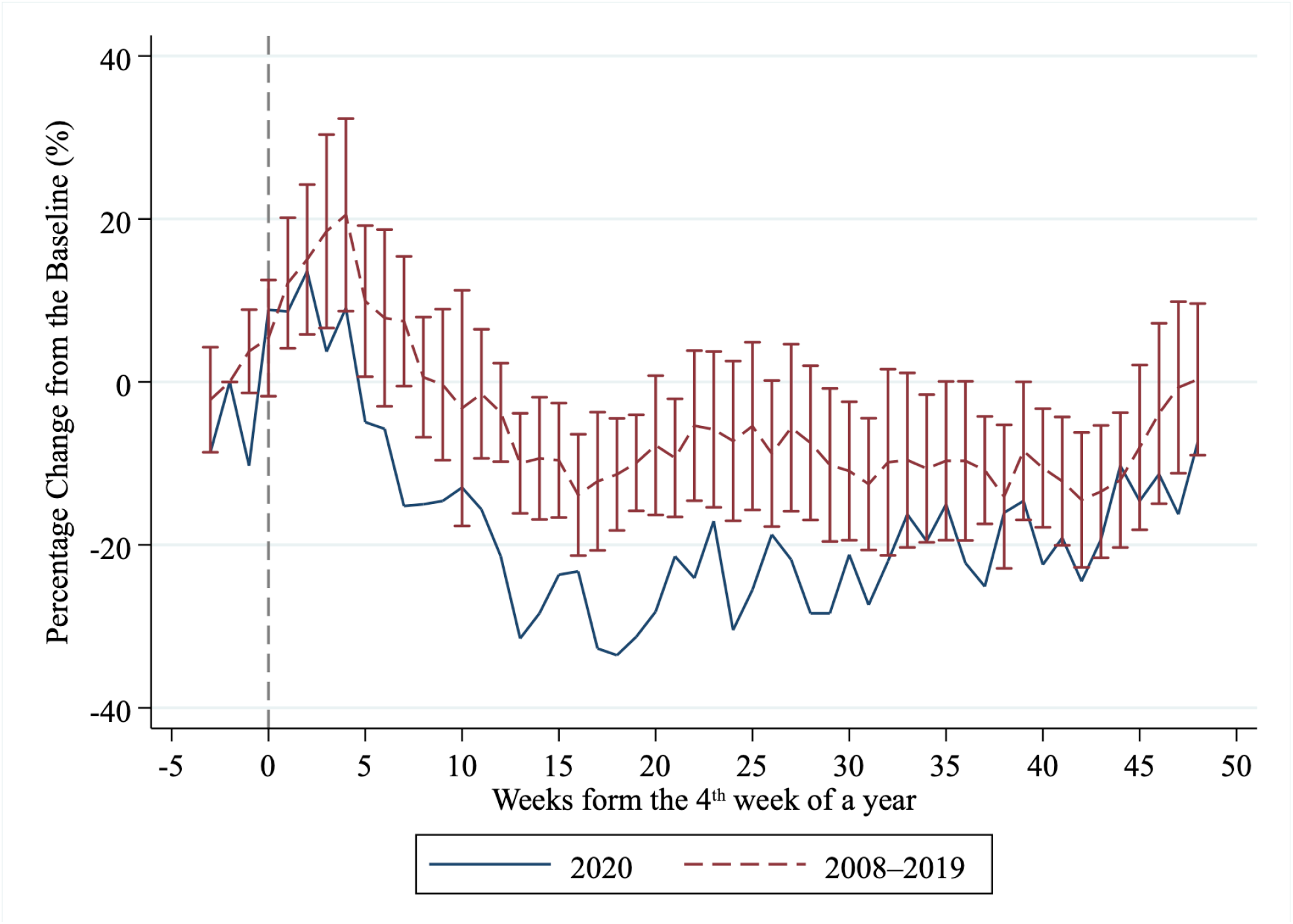
Effects of COVID-19 Outbreak on ILI Mortality. *Notes:* Figure 7 displays the percentage change in the weekly number of ILI-related deaths per 100,000 population from the baseline mean (i.e., the average outcome of the second weeks in each year). The solid line represents the trend of the ILI-related mortality rate in 2020. The dashed line represents the 12-year average of the ILI-related mortality rate during 2008–2019 and the corresponding 95% confidence interval (error bar). The vertical line in the graph denotes the 4^th^ week of a year.

### 5.2 Implications for the Observed Decline in Healthcare Utilization

Our results indicate that outpatient visits and inpatient admissions fell by 21% and 11%, respectively, during the pandemic period. For ILI diseases, the declines in outpatient visits and inpatient admissions were even larger (i.e. more than 50% decrease). Compared to estimates in recent studies, we find that the voluntary response is substantial. For example, Birkmeyer et al. (2020) found that hospital admissions for non-COVID-19 diseases decreased by more than 20% from February to April 2020 in the US. In addition, their results suggest that admissions for ILI diseases, such as urinary tract infection and pneumonia, fell by 40% to 50%.

Given the low risk of contracting COVID-19 in Taiwan, we believe our estimates could serve as a “lower bound” for voluntary healthcare utilization responses in other countries. This implies that we may treat voluntary behavior as a major reason for the observed decline in healthcare utilization. Furthermore, our results indicate that the demand for healthcare services did not get back to the pre-pandemic baseline, even after the pandemic died away in Taiwan, thereby suggesting that the COVID-19 outbreak might have had (and be having) a long-term impact on people’s health behaviors.

### 5.3 Implications for Healthcare Expenditure

Using our results, we can provide the estimated effect of the COVID-19 outbreak on NHI healthcare expenditure. Note that the average expense per outpatient visit and per inpatient admission are around 1,387 NT$ (i.e., 49.5 US$) and 63,249 NT$ (i.e., 2,258.8 US$), respectively. Based on the above information, our DID estimates suggest that the COVID-19 outbreak could have “saved” the NHI around 52.4 billion NT$ (i.e., 1.9 billion US$), which accounts for 7.3% of the annual NHI budget.^19^

## 6 Conclusion

This paper examines the effects of the COVID-19 outbreak on voluntary demand for healthcare in Taiwan. By comparing with the same period in the years before the COVID-19 outbreak (i.e. 2014–2019), we find a large decline in health utilization after the outbreak of COVID-19. On average, the number of outpatient visits and inpatient admissions decreased by 21% and 11% during the pandemic period, respectively. Even in the period with no local coronavirus cases, outpatient visits still remains a 9% reduction. In addition, the demand response of healthcare for ILI diseases was much larger and persistent than for non-ILI diseases. Finally, we also find that the outbreak induced a 10% to 30% decline in the ILI-mortality rate.

There are two important limitations of this paper. First, our data source only provides total outpatient visits/inpatient admissions and selected infectious diseases. Due to this limitation, we cannot analyze the impact of COVID-19 on other types of healthcare, such as mental health (Hamp-shire et al., 2021). It is possible that future researches will explore such issues after individual-level NHI claim data have been released. Second, our research design cannot untangle which prevention measures avoided the spread of ILI diseases and then reduced demand for associated healthcare. However, as universal masking has been the key element in Taiwan’s pandemic response, we believe that this practice should play an important role in this regard. Nevertheless, more studies are needed to understand the effect of different prevention measures on health utilization and health.

## Data Availability

Our healthcare utilization data originate from the Taiwan National ILI Disease Statistics System, accessed via the Taiwan Center for Disease Control's (TCDC) Open Data Portal. https://data.cdc.gov.tw/

## Online Appendix

### A Taiwan’s Response to the COVID-19 Pandemic

Taiwan had been praised by international medias as a success story of COVID-19 prevention.^20^ As of 9^th^ January 2021, Lowy Institute ranked Taiwan as the top three countries with the best performance on COVID-19 pandemic management.^21^ In general, three key strategies have helped the nation successfully prevent the spread of the virus: 1) Early border control and quarantine policies; 2) Distributing and producing face masks and 3) Disclosing COVID-19 information to the public.

#### A.1 Early Border Control and Quarantine Policies

As of the first confirmed case, the Taiwan government initiated quarantine policies requiring people returning form “high-risk” COVID-19 countries (e.g., China), and those who had come into contact with confirmed cases, had to enter self-quarantine for 14 days. From March 19^th^, the Taiwan government restricted all foreigners from entering the country, and on the very same day, all citizens returning from oversea had to take 14 days’ quarantine.^22^

#### A.2 Universal Use of Face Masks

In contrast to European and American countries, the Taiwan government considered face masks one of the most important items of personal protective equipment (PPE) for reducing COVID-19 transmission. In order to make sure every resident had access to face masks, at the beginning of the outbreak (i.e. January 24^th^, 2020) the Taiwan government banned their export and requisitioned a huge increase in local production. The daily production capacity of face mask manufacturers in Taiwan before the outbreak was 1.88 million pieces,^23^ but currently, Taiwan is able to produce more than 15 million per day.^24^ Moreover, starting from February 6^th^, 2020, the government implemented a name-based rationing system for face masks to curb panic-buying and to ensure the universal face-covering of all residents in Taiwan.

#### A.3 Public Disclosure of COVID-19 Information

Besides its universal masking policy, border controls and quarantine policies, Taiwan’s success in terms of controlling the epidemic can also be attributed to its information dissemination and disclosure strategies. On January 20^th^, 2020, the Central Epidemic Command Center (CECC) was initiated. When the first confirmed case was corroborated on January 22^nd^, the CECC held press conferences every day to report on the epidemic and to offer self-protection information to citizens.^25^

Specifically, the CECC reported newly confirmed cases, cumulative confirmed cases, new death cases and recovered cases every day. The CECC also set up an on-line system for citizens to find out daily data on COVID-19 cases relevant to different counties.^26^ In addition, when specific symptoms (such as loss of taste, stroke, etc.) were noted, the CECC also released this information to the public, and whenever any local cases were discovered, it highlighted these during the press conferences with particular emphasis on the source and route of infection. The above information made citizens aware of the severity of the epidemic and helped them monitor their personal health status carefully.^27^

#### A.4 Behavioral Responses to COVID-19 Information

Using Google Trends data, we find that the Taiwanese people responded to the announcement of the first confirmed case immediately by searching for information about the virus and personal protective equipment (PPE), such as face masks and sanitizer.^28^ Note that instead of showing absolute search volume, Google Trends only provides a relative measure for daily search volume ranging from 0–100, where the numbers represent the search volume relative to the highest point. A value of 100 is the peak popularity of the term, and a value of 50 means half as popular. In order to match the frequency of healthcare data, we aggregate daily data to the weekly level.

Figures A.1a suggests that the search intensity of the keywords “Coronavirus” nearly reached 250 in the week of the first confirmed case announcement.^29^ Moreover, this search intensity jumped more than double and reached its peak when the first local COVID-19 case was reported. We also find that PPE-related (i.e. face mask and sanitizer) searches also peaked after the announcement of the first local COVID-19 case (See Figure A.1c and A.1e).^30^ Due to the painful experience of the 2003 outbreak of SARS (Bennett et al., 2015), both the Taiwanese government and its people responded to the first COVID-19 case very quickly indeed.^31^

Not every country responded to the first COVID-19 case in such a way, with the United States being a counterexample. Consistent with Bento et al. (2020), Figures A.1b, A.1d and A.1f indicates that the information-seeking behavior of the American people was in fact immediate following the first COVID-19 case, which was reported on January 21^st^, 2020. However, in contrast to Taiwan, the peak of the relative search volume for COVID-19 and PPE-related key words happened on the 7^th^ week (i.e. March 8^th^ to 14^th^) after the first confirmed case, because the US government verified this case was COVID-19 on March 1^st^. In addition, the search intensity for face masks (see Figure A.1d) peaked after the Center for Disease Control and Prevention (CDC) recommended that people wearing a face mask (April 3^rd^) would be an effective way to prevent COVID-19 transmission on April 5^th^ to April 11^th^ (i.e. 11 weeks after first confirmed case).^32^ Figure A.2 shows the daily trends in search index, The patterns are similar to the weekly trends. Since the United States is a large country, it is possible that people only responded to local cases. In Figure A.3, we also find a similar pattern in search behavior, using Google Trends data for Washington State or Seattle, where the first COVID-19 cases happened.

According to a survey conducted by the National Taipei University of Nursing and Health Sciences in April,^33^ 97.5% of Taiwanese thought that coronavirus is a serious disease, and over 90% of the interviewees correctly answered questions regarding how the virus spreads and prevention measures. Figure A.4 in the Online Appendix A.4 shows the percentage of people who say they are wearing face mask when in public places across time, surveyed by YouGov (Smith, 2020). The figure shows that as early as in February, over 80% of Taiwanese said that they were wearing a face mask in a public space. Further, the portion of people wearing masks remains high through the time. In contrast, only 7% of Americans said they had worn a face mask in early March, and the proportion began raised to half only until mid-April. All of these results are consistent with the patterns seen in Google Trends data, suggesting that the Taiwanese people have responded to COVID-19 in a rapid and proactive way.

**Figure A.1:**
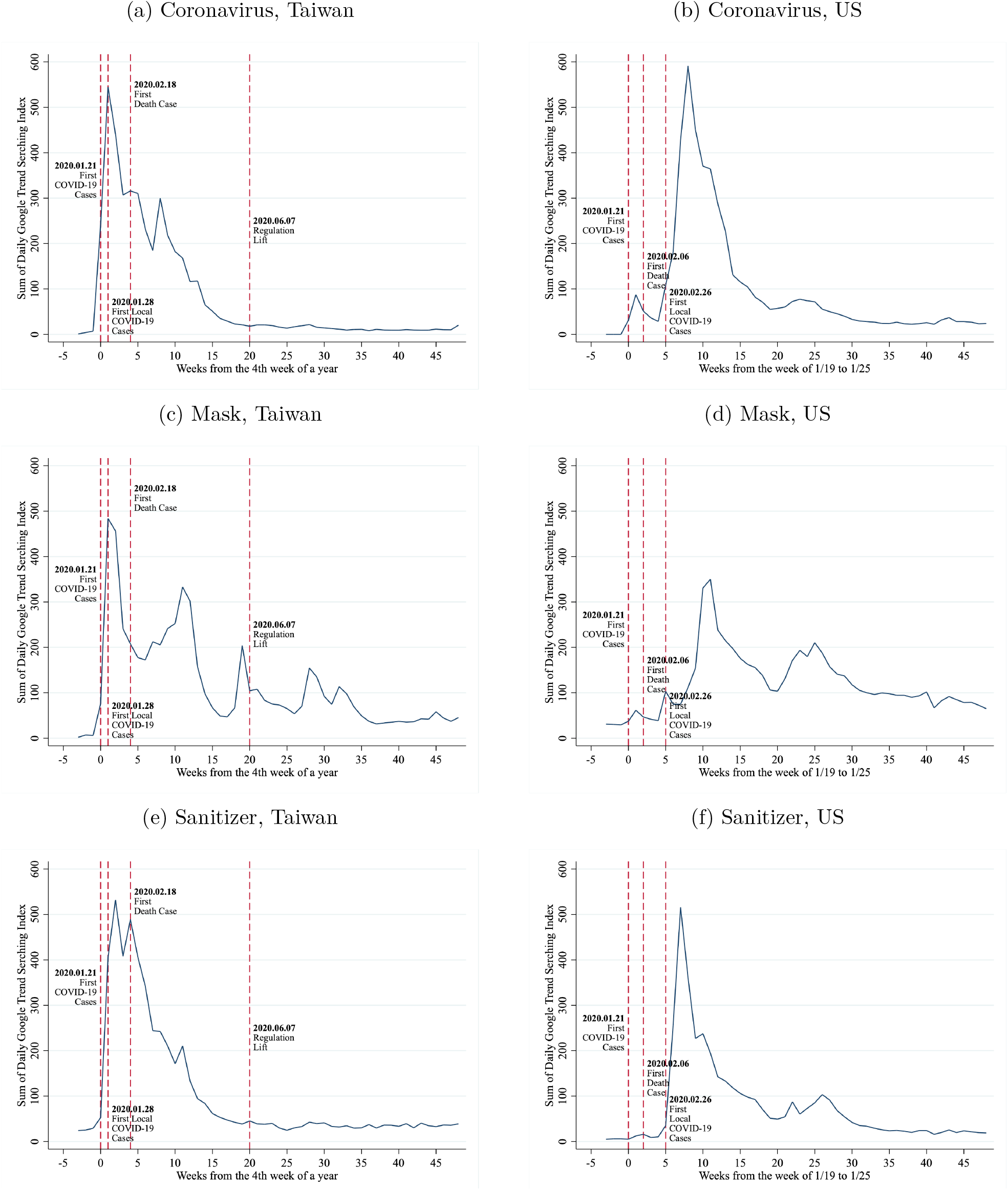
Google Search Intensity for COVID-19 Related Keywords: Taiwan and US. *Notes:* The figures are constructed by using Google Trends data. Google Trends only provides a relative measure for daily search volume ranging from 0–100. In order to match the frequency of healthcare data, we aggregate daily data to the weekly level. For Taiwan’s keywords, we use the equivalent term in Chinese for Coronavirus, mask, and sanitizer as the keywords.

**Figure A.2:**
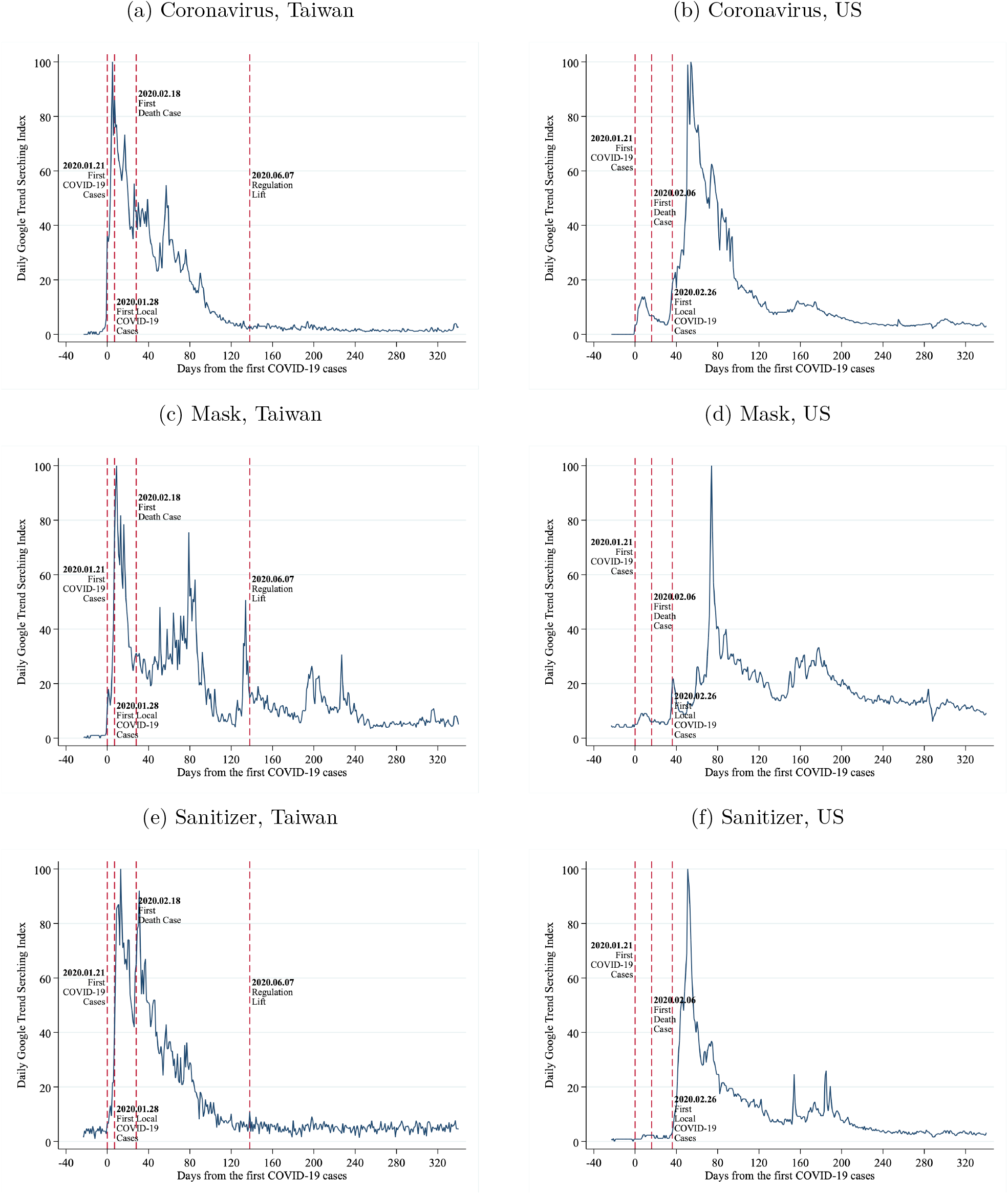
Google Search Intensity for COVID-19 Related Keywords: Taiwan and US. *Notes:* The figures are constructed by using Google Trends data. The daily search volume ranging from 0–100. For Taiwan’s keywords, we use the equivalent term in Chinese for Coronavirus, mask, and sanitizer as the keywords.

**Figure A.3:**
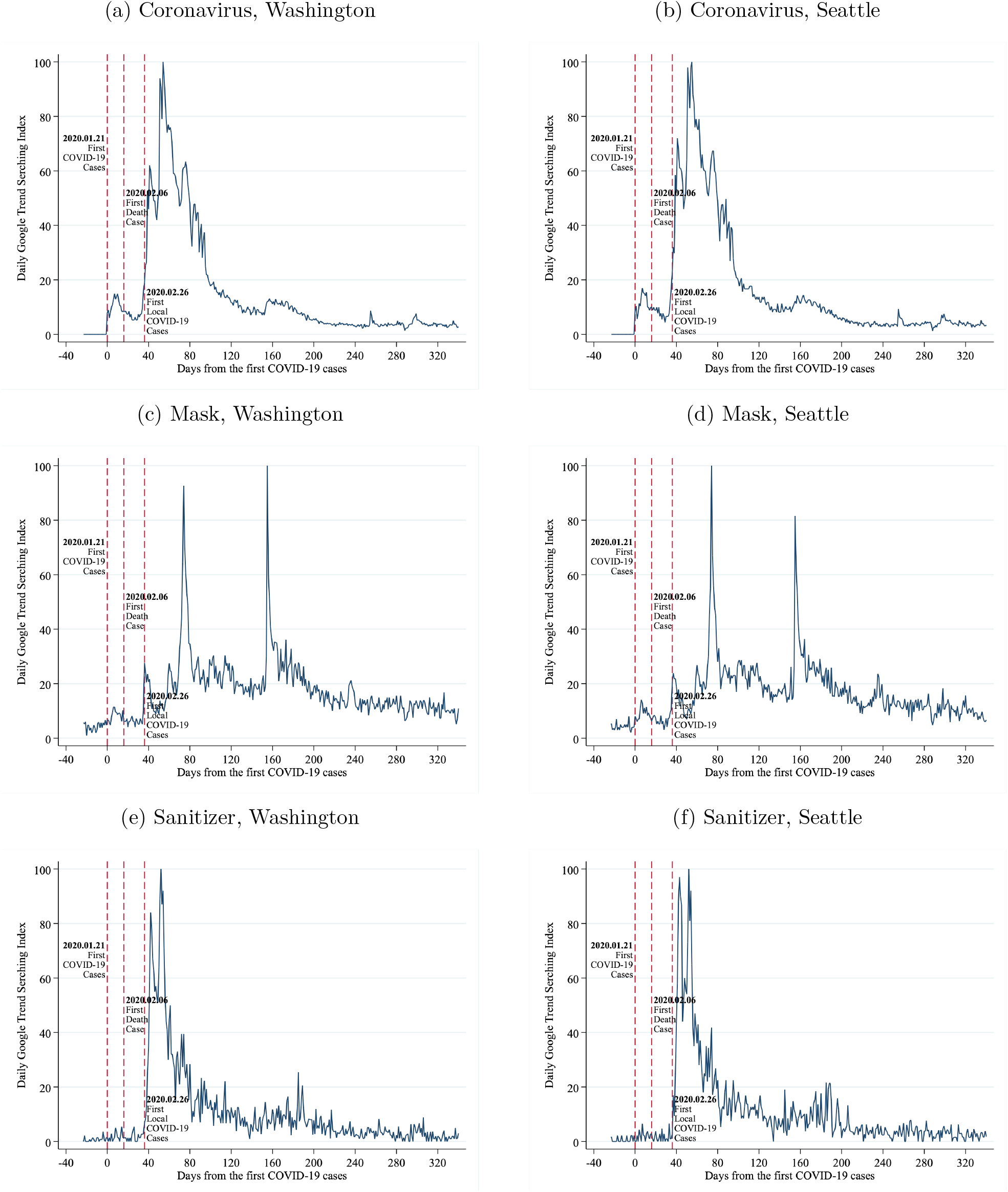
Google Search Intensity for COVID-19 Related Keywords: Washington and Seattle. *Notes:* The figures are constructed by using Google Trends data. Google Trends only provides a relative measure for daily search volume ranging from 0–100.

**Figure A.4:**
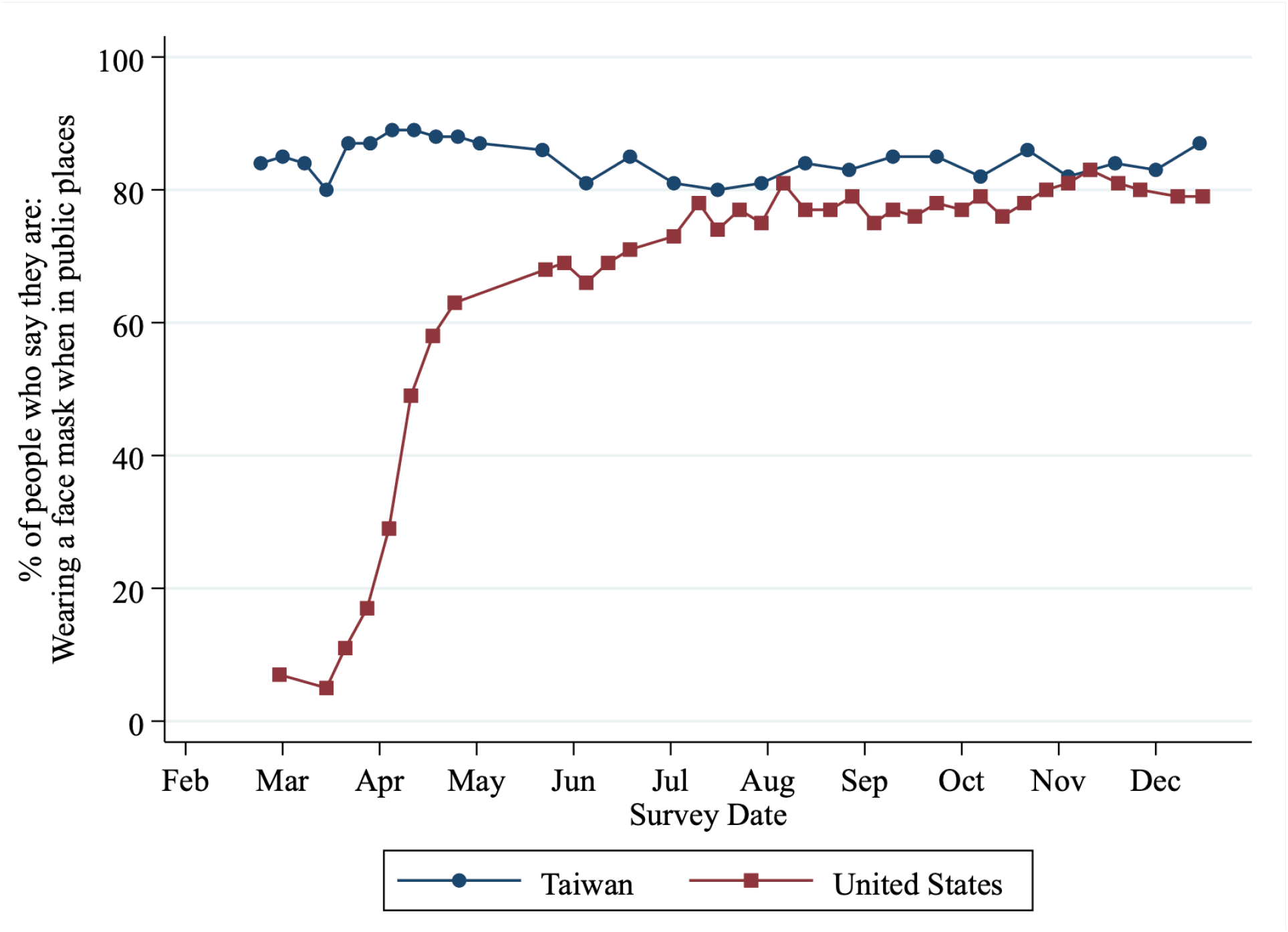
Share of People Wearing a Face Mask When in Public Space. *Notes:* Data source is from YouGov (Smith, 2020)

### B Additional Tables

**Table B.1:** Robustness Check: Clustering levels of Standard Errors (DID Design)

|  | (1) | (2) | (3) | (4) | (5) | (6) | (7) | (8) |
| --- | --- | --- | --- | --- | --- | --- | --- | --- |
|  | Outpatient Care |  |  |  | Inpatient Care |  |  |  |
| **Panel A:** All diseases |  |  |  |  |  |  |  |  |
| $Y_{2020} \times Post$ | -0.14*** | -0.13*** | -0.14*** | -0.14*** | -0.05*** | -0.04*** | -0.05 | -0.04 |
| Cluster at County Level | [0.01] | [0.01] | [0.01] | [0.01] | [0.03] | [0.03] | [0.03] | [0.03] |
| Cluster at YearWeek Level | {0.02} | {0.02} | 0.00} | {0.02} | {0.02} | {0.02} | {0.02} | {0.02} |
| **Panel B:** ILI diseases |  |  |  |  |  |  |  |  |
| $Y_{2020} \times Post$ | -0.46*** | -0.47*** | -0.47*** | -0.48*** | -0.51*** | -0.52*** | -0.51*** | -0.50*** |
| Cluster at County Level | [0.01] | [0.02] | [0.01] | [0.01] | [0.05] | [0.05] | [0.05] | [0.05] |
| Cluster at YearWeek Level | {0.04} | {0.04} | 0.00} | {0.04} | {0.05} | {0.05} | {0.05} | {0.05} |
| **Panel C:** Non-ILI diseases |  |  |  |  |  |  |  |  |
| $Y_{2020} \times Post$ | -0.10*** | -0.10*** | -0.11*** | -0.11*** | -0.02*** | -0.01 | -0.02 | -0.02 |
| Cluster at County Level | [0.01] | [0.01] | [0.01] | [0.01] | [0.03] | [0.03] | [0.03] | [0.03] |
| Cluster at YearWeek Level | {0.02} | {0.02} | {0.02} | {0.02} | {0.02} | {0.02} | {0.02} | {0.02} |
| Observation | 8,008 |  |  |  |  |  |  |  |
| Basic control | ✓ | ✓ | ✓ | ✓ | ✓ | ✓ | ✓ | ✓ |
| Demographic variables |  | ✓ | ✓ |  |  | ✓ | ✓ |  |
| Weather variables |  | ✓ | ✓ | ✓ |  | ✓ | ✓ | ✓ |
| County fixed effect |  |  | ✓ | ✓ |  |  | ✓ | ✓ |
| County-by-year fixed effect |  |  |  | ✓ |  |  |  | ✓ |
| County-by-week fixed effect |  |  |  | ✓ |  |  |  | ✓ |
| County specific time trend |  |  |  | ✓ |  |  |  | ✓ |

**Table B.2:**
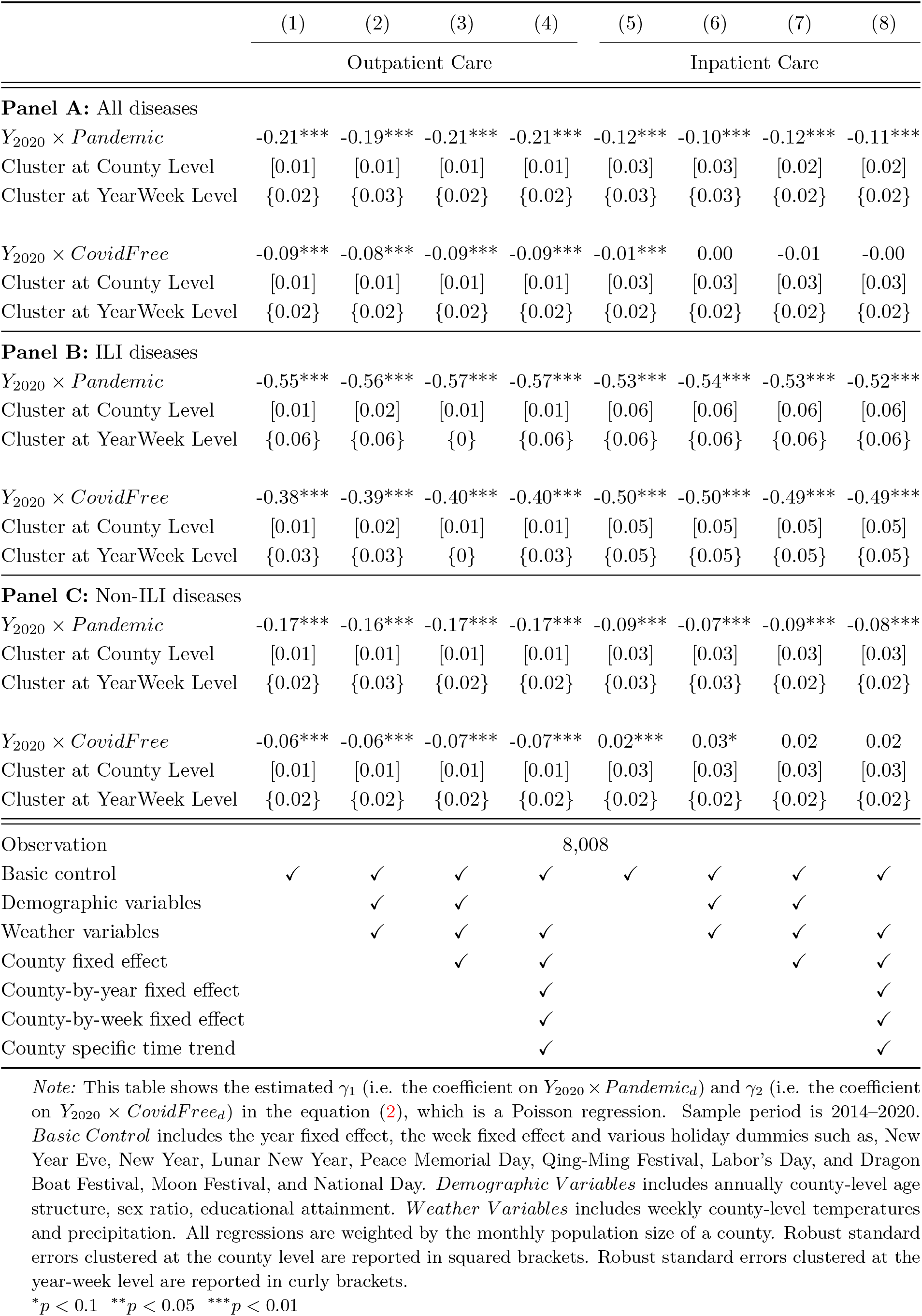
Robustness Check: Clustering levels of Standard Errors (Multi-period DID Design)

**Table B.3:** Robustness Check: Unweighted Regressions (DID Design)

|  | (1) | (2) | (3) | (4) | (5) | (6) | (7) | (8) |
| --- | --- | --- | --- | --- | --- | --- | --- | --- |
|  | Outpatient Care |  |  |  | Inpatient Care |  |  |  |
| **Panel A:** All diseases |  |  |  |  |  |  |  |  |
| $Y_{2020} \times Post$ | -0.13***<br>(0.00) | -0.11***<br>(0.02) | -0.13***<br>(0.02) | -0.13***<br>(0.02) | -0.05***<br>(0.00) | -0.05*<br>(0.03) | -0.05<br>(0.03) | -0.05<br>(0.03) |
| **Panel B:** ILI diseases |  |  |  |  |  |  |  |  |
| $Y_{2020} \times Post$ | -0.43***<br>(0.03) | -0.43***<br>(0.04) | -0.45***<br>(0.04) | -0.45***<br>(0.04) | -0.56***<br>(0.09) | -0.56***<br>(0.09) | -0.57***<br>(0.11) | -0.57***<br>(0.11) |
| **Panel C:** Non-ILI diseases |  |  |  |  |  |  |  |  |
| $Y_{2020} \times Post$ | -0.10***<br>(0.00) | -0.08***<br>(0.02) | -0.10***<br>(0.02) | -0.10***<br>(0.02) | -0.02***<br>(0.00) | -0.01<br>(0.03) | -0.02<br>(0.03) | -0.01<br>(0.03) |
| Observation | 8,008 |  |  |  |  |  |  |  |
| Basic control | ✓ | ✓ | ✓ | ✓ | ✓ | ✓ | ✓ | ✓ |
| Demographic variables |  | ✓ | ✓ |  |  | ✓ | ✓ |  |
| Weather variables |  | ✓ | ✓ | ✓ |  | ✓ | ✓ | ✓ |
| County fixed effect |  |  | ✓ | ✓ |  |  | ✓ | ✓ |
| County-by-year fixed effect |  |  |  | ✓ |  |  |  | ✓ |
| County-by-week fixed effect |  |  |  | ✓ |  |  |  | ✓ |
| County specific time trend |  |  |  | ✓ |  |  |  | ✓ |

**Table B.4:**
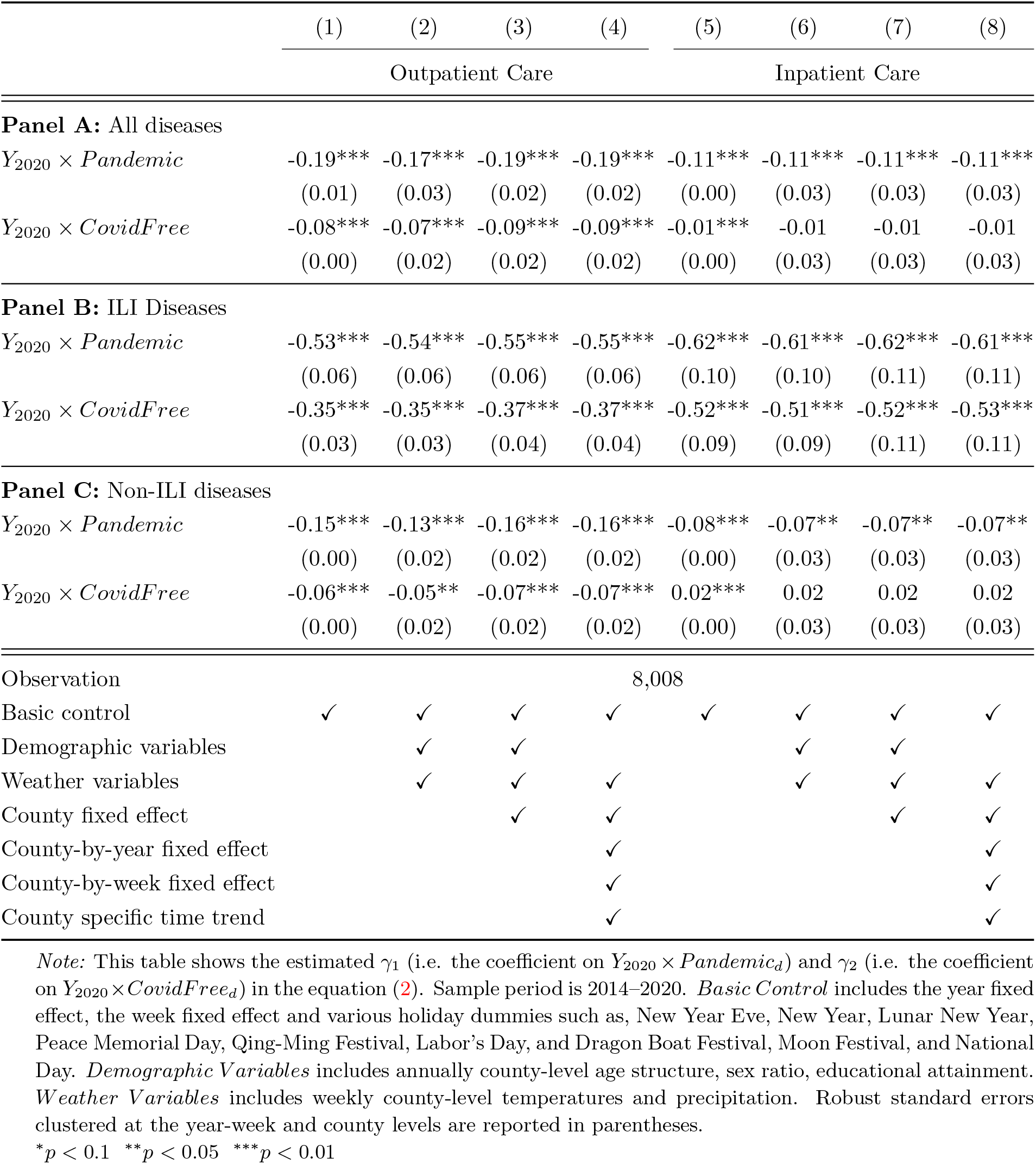
Robustness Check: Unweighted Regressions (Multi-period DID Design)

|  | (1) | (2) | (3) | (4) | (5) | (6) | (7) | (8) |
| --- | --- | --- | --- | --- | --- | --- | --- | --- |
|  | Outpatient Care |  |  |  | Inpatient Care |  |  |  |
| **Panel A: All diseases** |  |  |  |  |  |  |  |  |
| $Y_{2020} \times Pandemic$ | -0.19***<br>(0.01) | -0.17***<br>(0.03) | -0.19***<br>(0.02) | -0.19***<br>(0.02) | -0.11***<br>(0.00) | -0.11***<br>(0.03) | -0.11***<br>(0.03) | -0.11***<br>(0.03) |
| $Y_{2020} \times CovidFree$ | -0.08***<br>(0.00) | -0.07***<br>(0.02) | -0.09***<br>(0.02) | -0.09***<br>(0.02) | -0.01***<br>(0.00) | -0.01<br>(0.03) | -0.01<br>(0.03) | -0.01<br>(0.03) |
| **Panel B: ILI Diseases** |  |  |  |  |  |  |  |  |
| $Y_{2020} \times Pandemic$ | -0.53***<br>(0.06) | -0.54***<br>(0.06) | -0.55***<br>(0.06) | -0.55***<br>(0.06) | -0.62***<br>(0.10) | -0.61***<br>(0.10) | -0.62***<br>(0.11) | -0.61***<br>(0.11) |
| $Y_{2020} \times CovidFree$ | -0.35***<br>(0.03) | -0.35***<br>(0.03) | -0.37***<br>(0.04) | -0.37***<br>(0.04) | -0.52***<br>(0.09) | -0.51***<br>(0.09) | -0.52***<br>(0.11) | -0.53***<br>(0.11) |
| **Panel C: Non-ILI diseases** |  |  |  |  |  |  |  |  |
| $Y_{2020} \times Pandemic$ | -0.15***<br>(0.00) | -0.13***<br>(0.02) | -0.16***<br>(0.02) | -0.16***<br>(0.02) | -0.08***<br>(0.00) | -0.07**<br>(0.03) | -0.07**<br>(0.03) | -0.07**<br>(0.03) |
| $Y_{2020} \times CovidFree$ | -0.06***<br>(0.00) | -0.05**<br>(0.02) | -0.07***<br>(0.02) | -0.07***<br>(0.02) | 0.02***<br>(0.00) | 0.02<br>(0.03) | 0.02<br>(0.03) | 0.02<br>(0.03) |
| Observation | 8,008 |  |  |  |  |  |  |  |
| Basic control | ✓ | ✓ | ✓ | ✓ | ✓ | ✓ | ✓ | ✓ |
| Demographic variables |  | ✓ | ✓ |  |  | ✓ | ✓ |  |
| Weather variables |  | ✓ | ✓ | ✓ |  | ✓ | ✓ | ✓ |
| County fixed effect |  |  | ✓ | ✓ |  |  | ✓ | ✓ |
| County-by-year fixed effect |  |  |  | ✓ |  |  |  | ✓ |
| County-by-week fixed effect |  |  |  | ✓ |  |  |  | ✓ |
| County specific time trend |  |  |  | ✓ |  |  |  | ✓ |

## Footnotes

1 Data source: Our World In Data. https://ourworldindata.org/coronavirus. Date accessed: September 6^th^, 2021

2 Recent studies indicate that the pandemic and lockdown policies have led to a large decrease in outpatient visits (Baum et al., 2021; Borrelli et al., 2020; Chatterji and Li, 2021; Dewar et al., 2020; Kasle et al., 2020; Kutlu et al., 2020; Mehrotra et al., 2020; Patel et al., 2021; Wosik et al., 2021; Ziedan et al., 2020), emergency visits (Giannouchos et al., 2021; Hartnett et al., 2020; Holland et al., 2021; Lange et al., 2020; Lerner et al., 2020), and inpatient admissions (Abbas et al., 2021; Birkmeyer et al., 2020; Gómez-Ramiro et al., 2021; Pelletier et al., 2021). Most studies focus on the US, although a few have been conducted on the UK, Turkey, Spain, and Italy; however, none of them has examined or compared the effects on outpatient and inpatient departments simultaneously. Besides, most of these studies only use data from single medical institutions, single health insurance providers, or data sources with a limited number of hospitals/clinics.

3 The Urban Institute Coronavirus Tracking Survey indicates that one-third of adults report encountered an “unmet need for medical care,” due to COVID-19 (Gonzalez et al., 2020). Furthermore, the KFF Health Tracking Poll reveals that during the COVID-19 outbreak, half of the US population skipped or postponed medical care, even though onethird of them did try to access medical treatment over a 3-month period (Hamel et al., 2020).

4 Since such an achievement was highly unique during 2020, several news outlets reported this record, for example *Time Magazine* (see https://time.com/5905129/taiwan-coronavirus-record/)

5 The COVID-19 pandemic was well controlled in Taiwan during the first half of 2021, with daily cases remaining below 30 till mid-May (most cases were imported and number of daily local cases was zero). During May 15^th^ to the end of June, 2021, Taiwan experienced a second-wave outbreak of local transmissions, nevertheless daily new confirmed cases remained at around 10 to 25 per million people, which is still a lower bound compared to other nations. In August, Taiwan only had daily COVID-19 cases fewer than 20. On August 25^th^, Taiwan had no COVID-19 case. Note that we do not include the 2021 period in our analysis.

6 For example, Ziedan et al. (2020) analyzes how state closure policy affected non-COVID-19 health utilization. The result suggests that through March and early April, outpatient visits in the US declined by around 40%, and one-third of this reduction (i.e., a 15% decline in outpatient visits) could be explained by states’ closure policies. Birkmeyer et al. (2020) examine changes in non-COVID-19 hospital admissions after the COVID-19 outbreak. They find out that hospitals seriously affected by COVID-19 experienced a larger reduction in non-COVID-19 admissions than those with a minimal shock from the outbreak.

7 This study is also related to the body of theoretical and empirical literature on voluntary avoidance behavior (Perra et al., 2011; Funk et al., 2010; Rubin et al., 2009; Bayham et al., 2015). For example, several recent studies have found that people’s voluntary response plays an important role in what decisions they make in terms of mobility, social distancing, and mask-wearing during the pandemic (Gupta et al., 2020; Yan et al., 2021; Chudik et al., 2020; Farboodi et al., 2020; Allcott et al., 2020; Herby, 2021). We contribute to this stream of the literature by quantifying the voluntary avoidance of healthcare utilization during the pandemic.

8 As of the end of 2020, the incidence of COVID-19 per million population was 33.55 in Taiwan and 60,723 in the US. Sources: https://ourworldindata.org/covid-cases. Date accessed: May. 29^th^, 2021

9 The government also suspended some of the sex-related leisure and entertainment venues from April 9^th^.

10 https://data.cdc.gov.tw/

11 https://www.ris.gov.tw/app/portal/346

12 All demographic variables are measured at annual level. The variables for age structure are share of population aged 0–14, 15–64, and above 65. The variable for sex ratio is the ratio of females to males in a population. The variables for educational attainment are share of population who hold post-secondary degree, high-school degree, and no high-school degree.

13 https://statis.moi.gov.tw/micst/stmain.jsp?sys=100

14 https://e-service.cwb.gov.tw/HistoryDataQuery/index.jsp

15 Some years have a 53^th^ week, but we do not include that period.

16 ILI diseases include influenza, non-COVID-19 pneumonia, and acute upper respiratory infections.

17 The time trend is constructed as the number of weeks from the first week of 2014.

18 Note that we do not include demographic variables, which are measured at county-by-year level, when controlling county-by-year fixed effects.

19 We use estimates in column (4) and (8) in Panel A of Table 3 and number of total outpatient visits (inpatient admissions) in pre-outbreak period to obtain the change in healthcare expenditure induced by COVID-19 pandemic (around 52.4 billion NT$). Compared to the NHI annual budget in 2019, which was 715.3 billion NT$, our result suggests that COVID-19 outbreak reduced annual healthcare budget by 7.3%.

20 Including Reuters, New York Time, CNN and Economist (https://www.reuters.com/article/uk-gay-pride-taiwan-idUKKBN27G0CM, https://www.reuters.com/article/us-health-coronavirus-taiwan-idUKKBN27Y0R7, https://www.nytimes.com/2021/03/13/world/asia/taiwan-covid.html, https://www.nytimes.com/2021/03/13/world/taiwan-a-covid-19-outlier-is-selling-something-scarce-life-without-fear-of-the-virus.html, https://www.cnn.com/2020/10/29/asia/taiwan-covid-19-intl-hnk, https://www.cnn.com/2020/04/04/asia/taiwan-coronavirus-response-who-intl-hnk/index.html, https://www.economist.com/asia/2020/12/02/covid-19-has-ravaged-economies-all-over-the-world-but-not-taiwans. Date Accessed: May 26^th^, 2021)

21 The first place is New Zealand, and the second place is Vietnam. Totally 116 countries were evaluated. https://interactives.lowyinstitute.org/features/covid-performance. Date Accessed: May 26^th^, 2021.

22 The border control policy for the foreigners had been relaxed from June 29^th^. But foreigners entry from other countries need to provide COVID-19 testing with negative results.

23 https://www.moea.gov.tw/MNS/populace/news/News.aspx?kind=1&menu_id=40&news_id=88545, Date accessed: Sep. 5^th^, 2020

24 https://www.moea.gov.tw/MNS/populace/news/News.aspx?kind=9&menu_id=22333&news_id=89290, Date accessed: Sep. 5^th^, 2020

25 The frequency of these briefings was reduced to once a week from June 8^th^, 2020, following a consecutive 8 weeks of no local confirmed cases.

26 https://nidss.cdc.gov.tw/ch/NIDSS_DiseaseMap.aspx?dc=1&dt=5&disease=19CoV

27 More detailed review on Taiwan’s pandemic responses can be found in Wang (2020).

28 Google Trends, powered by Google, provides the relative search interests of a given keyword made to Google at a given time period and location. Readers can get data from this website: https://trends.google.com.tw/.

29 We use the equivalent term in Chinese for Coronavirus as the keywords.

30 We use the equivalent term in Chinese for mask and sanitizer as the keywords.

31 SARS severely hurt Taiwan in 2003, with a total of 668 reported cases and 181 death cases (Chen et al., 2005).

32 https://www.livescience.com/cdc-recommends-face-masks-coronavirus.html, Date accessed: Aug. 20^th^, 2020

33 https://news.sina.com.tw/article/20200509/35111198.html, Date accessed: Aug. 10^th^, 2020

